# Development, testing and validation of a SARS-CoV-2 multiplex panel for detection of the five major variants of concern on a portable PCR platform

**DOI:** 10.1101/2022.08.02.22277351

**Authors:** Bryce J Stanhope, Brittany Peterson, Brittany Knight, Ray Nobles Decadiz, Roger Pan, Phillip Davis, Anne Fraser, Manunya Nuth, Jesse vanWestrienen, Erik Wendlandt, Bruce Goodwin, Chris Myers, Jennifer Stone, Shanmuga Sozhamannan

## Abstract

Many SARS-CoV-2 variants have emerged during the course of the COVID-19 pandemic. These variants have acquired mutations conferring phenotypes such as increased transmissibility or virulence, or causing diagnostic, therapeutic, or immune escape. Detection of Alpha and the majority of Omicron sublineages by PCR relied on the so-called S gene target failure due to the deletion of six nucleotides coding for amino acids 69-70 in the spike (S) protein. Detection of hallmark mutations in other variants present in samples relied on whole genome sequencing. However, whole genome sequencing as a diagnostic tool is still in its infancy due to geographic inequities in sequencing capabilities, higher cost compared to other molecular assays, longer turnaround time from sample to result, and technical challenges associated with producing complete genome sequences from samples that have low viral load and/or high background. Hence, there is a need for rapid genotyping assays. In order to rapidly generate information on the presence of a variant in a given sample, we have created a panel of four triplex RT-qPCR assays targeting 12 mutations to detect and differentiate all five variants of concern: Alpha, Beta, Gamma, Delta and Omicron. We also developed an expanded pentaplex assay that can reliably distinguish among the major sublineages (BA.1-BA.5) of Omicron. *In silico*, analytical and clinical testing of the variant panel indicate that the assays overall exhibit high sensitivity and specificity. This variant panel can be used as a Research Use Only screening tool for triaging SARS-CoV-2 positive samples prior to whole genome sequencing.

## BACKGROUND

The emergence of COVID-19 in late 2019 in Wuhan, China and subsequent spread of the virus across the globe led to the unprecedented COVID-19 pandemic and declaration as a Public Health Emergency of Global Concern on Jan 30, 2020 by the World Health Organization (WHO) (1). Identification of the etiological agent of COVID-19 as SARS-CoV-2 and release of the first whole genome sequence of SARS-CoV-2 shortly thereafter on Jan 11, 2020 have been instrumental in the rapid development of diagnostics, therapeutics and vaccines (2, 3). Public health measures such as ‘test, trace and isolate’ and individual health measures, such as treating symptomatic individuals as needed, rely on rapid and accurate diagnosis of infection early in the course of the infection cycle. The first molecular diagnostic test was issued an emergency use authorization (EUA) on Feb 04, 2020. In the last two years, as of May 05, 2022, there are 307 EUA SARS-CoV-2 molecular tests and 48 antigen tests (4). COVID-19 diagnosis is further complicated by the fact that as much as 40% of infections may be asymptomatic depending on the population cohorts, where secondary transmission might still be occurring (5). Hence, PCR tests remain to be the gold standard for SARS-CoV-2 detection because of their exquisite sensitivity and better performance identifying asymptomatic cases (6). It has been argued that PCR tests may show positivity for a longer duration even after the infectivity has subsided and hence rapid antigen tests may be more reflective of infectivity (7, 8). However, PCR tests are preferred especially in testing asymptomatic infections and early in the infection cycle (9, 10).

Despite the availability of hundreds of different tests, testing demands and needs have not been fully met and there is always a shortage of test quantities and sites around the globe. This in turn affect rapid decision making to test, trace and isolate infected persons to prevent secondary transmissions (11). For this reason, there is a need for rapid and accurate tests that have higher sensitivity and can be performed in less complex settings. Indeed, at-home diagnostic kits meet the turn-around time requirements, but few options are currently available for sensitive PCR-based tests compared to the less sensitive rapid antigen tests (12). In addition to testing constraints, the emergence of variants has further complicated testing capabilities.

Rapid proliferation and sustained transmission of SARS CoV-2 in the human population have led to waves of variants that swept through the globe (13, 14). To identify the variants, genomic surveillance has come to the forefront of COVID-19 pandemics as evidenced by the tremendous explosion of whole genome sequencing data from all around the globe (15). Whole genome sequencing has become ubiquitous and as of May 24, 2022, more than 11 million sequences have been generated and deposited by the global sequencing community to the GISAID database (16-18). Whole genome sequence data have been extensively used in determining the changing genetic profiles of viral variants and tracking viral mobility across communities using the genetic changes, in what has come to be known as genomic epidemiology (19-21). Rapid identification of the prevalent variants without having to wait weeks for sequence data is paramount for public health measures and decision-making and in some instances even treatment options. For example, some antibody therapies fail against Omicron infections due to specific mutations in critical positions in the spike protein (22). Hence, there is a need for genotyping PCR assays that have rapid turn-around times from sample to results on par with other PCR diagnostic tests (hours instead of days) compared to 14 days or more for sequencing results (23-33).

Here, we describe a panel of multiplex RT-qPCR assays to detect and differentiate all five WHO variants of concern: Alpha, Beta, Gamma, Delta and Omicron. These RT-qPCR assays can be performed on a hand-held, battery operated device, the Biomeme Franklin™ three9 Real-Time PCR Thermocycler. We chose the Biomeme platform because it is portable, easy to use, battery-powered, and hence, field deployable with a minimal footprint. Biomeme also provides multiple assay formats—including lyophilized, room-temperature stable 3-well Go-Strips and 96-well Go-Plates—that can be used in both point-of-need, low-throughput devices like the Franklin™, as well as in traditional lab-based, high-throughput instruments with multiplex capabilities.

## MATERIALS AND METHODS

### Primers, probes and PCR reagents

The primer/probe and template sequences that were designed in this study are presented in Table S1 and Table S2 respectively. Reagents and assay conditions for various assay formats are described under different subsections below.

### Biomeme Assay design, testing and validation workflow

### Assay design for Triplexes 1-3

The whole genomes of SARS-CoV-2 variants were downloaded from the GISAID database and aligned to the genome of the Wuhan-Hu-1 isolate (GenBank accession no. MN908947.3) using Clustal Omega (https://www.ebi.ac.uk/Tools/msa/clustalo/). Primers were designed at conserved regions of the spike (S) and nucleocapsid (N) genes to generate amplicons comprising the mutations of interest using the Primer Quest Software (https://www.idtdna.com/PrimerQuest/Home/Index) from Integrated DNA Technologies, Inc. (IDT; Coralville, Iowa, USA) and analyzed for potential dimerization and hairpin products by the OligoAnalyzer tool (IDT). Probes containing LNA and conjugated minor groove binder (MGB) were manually designed. In particular, the assistance of IDT’s in-house tools afforded the predication of melting temperatures and mismatch discrimination potentials. The S:Y144del primer and probe sequences were designed according to Vogels et al. (34). Primer candidates were then analyzed by Primer-BLAST (https://www.ncbi.nlm.nih.gov/tools/primer-blast/) and the probes by blastn (https://blast.ncbi.nlm.nih.gov/Blast.cgi?PAGE_TYPE=BlastSearch) to ensure the recognition of the intended targets. For suitability of multiplexing, primer combinations were predicted by MultiPLX 2.1 (35). Primers and LNA probes were purchased either from IDT or Sigma-Aldrich (St. Louis, MO, USA), with the primers purified by standard desalting and the probes by HPLC. The MGB probe was purchased from Applied Biosystems (Foster City, CA, USA) as HPLC-purified products. gBlocks were purchased from IDT.

### RT-qPCR on Standard Thermocyclers

One-step reverse transcription-quantitative polymerase chain reaction (RT-qPCR) was performed to avoid potential cross-contamination. A typical 20-µL reaction containing the DNA or RNA sample was prepared in LyoRNA 2.0 Master Mix (Biomeme, Inc., Philadelphia, PA, USA) according to the manufacturer’s instructions and assayed on a Bio-Rad CFX96 instrument (Bio-Rad Laboratories, Inc., Hercules, CA, USA) in a triplex mixture. The assays were run under the following conditions: reverse transcription at 55°C for 2 minutes, initial denaturation at 95°C for 1 minute, and 45 cycles of 95°C for 3 seconds and annealing/extension at 62°C for 30 seconds. Assay development was accomplished using gBlock templates in the RT-qPCR reactions and later confirmed with *in vitro*-transcribed RNA templates generated by the MEGAscript T7 Transcription Kit (Invitrogen, Waltham, MA, USA) according to the manufacturer’s instructions using 80 ng of gBlock templates. Triplexes were tested against both wild-type and mutant templates, and only the probes that could specifically discriminate between the two templates were selected for optimization.

### RT-qPCR on the Biomeme Franklin™ Thermocycler

Incorporating the same primer and probe concentrations used for measurement on the Bio-Rad CFX96 instrument, triplex assays were assembled into freeze-dried test strips (Go-Strips and Go-Plates) using Biomeme’s proprietary formulations. Each well of the Go-Strip or Go-Plate was reconstituted with 20 μL total volume of nuclease-free water and template DNA/RNA. The Go-Strips were then loaded into the portable Franklin™ Real-Time Thermocycler (36) in the appropriate orientation and monitored by the Biomeme software application on the included smartphone, while the Go-Plates were measured on the Bio-Rad CFX96 instrument. RT and cycling conditions used on all platforms were the same as those described for the LyoRNA master mix.

### Lyophilized Assay Manufacturing

Triplex assays were lyophilized into Go-Strip and Go-Plate formats. Go-Strips contained three reactions packaged individually in a small foil pouch designed for use on the Biomeme Franklin™. Go-Plates are 96-well plates packaged in a foil pouch meant for standard laboratory RT-qPCR systems. Go-Strips and Go-Plates are shelf-stable at room temperatures for 18 months in their original packaging.

### Omicron assay design and optimization

Probes for the omicron multiplexes (Triplex 4 and the Omicron pentaplex) were manually designed to overlie the targeted mutation site, using Primer Express® Software Version 3.0.1 to estimate melting temperatures for both the standard and MGB probes. Candidate primer pairs were designed using Primer3 Plus (37), then checked against the NCBI nt database for off-target matches and on-target mismatches using the NCBI Primer-BLAST tool (38). Candidate primer pairs that passed NCBI Primer-BLAST screening and candidate probe sequences were then tested for multiplex compatibility (oligo interactions and melting temperature) using ThermoFisher’s Multiple Primer Analyzer (39), and some manual adjustments to primer sequences were made to enhance compatibility. Candidate primer and probe combinations were then screened, down-selected, and optimized using 10-fold serial dilutions of template (50 to 50,000 copies per reaction) to evaluate sensitivity, efficiency, and signal-to-noise ratios.

Candidate assays were also tested against a high-level of wild-type genomic RNA to confirm exclusivity against non-variant template. Initial testing and optimization were performed in singleplex reactions using gBlock templates, but then progressed to multiplex testing using synthetic SARS-CoV-2 gRNA commercially available from Twist Biosciences (San Francisco, CA, USA), as well as RNA extracted from patient specimens and stock cultures (once available). Because gRNA could not be obtained for BA.1.1, BA.3, BA.4, and BA.5, gBlocks encompassing the S gene and relevant portion of the N gene were designed based on reference sequences, and serial dilutions of those were used for testing.

### Verification of assay signatures *in silico*

To evaluate inclusivity of each assay on a per-lineage basis, SARS-CoV-2 genomes designated as ‘complete’ (N=1005125) were downloaded from NCBI Virus (DOI: 10.1093/nar/gkw1065) on March 31, 2022, along with a corresponding metadata table containing the Pangolin (https://doi.org/10.1038/s41564-020-0770-5) lineage assignments. Upon observing that BA.3 lineage coverage was limited to a single record in the dataset drawn from NCBI Virus, we supplemented the dataset by downloading BA.3 assigned records (N=4370) from GISAID.org on April 15, 2022. Similarly, BA.4 sequences (N=1174) and BA.5 sequences (N=663) were downloaded from GISAID.org on May 11, 2022. A complete table of author attributions for the GISAID sequences is available in Supplementary Table S3.

Inclusivity was calculated as set coverage for each assay target for each lineage. A lineage’s coverage was calculated by dividing the *in silico* predicted PCR-positive sequences (based on cutoff criteria for primer and probe alignment mismatches) by the total number of sequences assigned that lineage in the database. Formally, the inclusivity *C* of each primer set is: 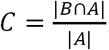, where *B* is the set of records returned by a primer pair/probe and *A* is the set of records for a given lineage.

*In silico* prediction of amplicon products for the evaluated assays was conducted using a modified version of simulate_PCR (https://doi.org/10.1186/1471-2105-15-237) re-implemented in Python. This approach relies on BLAST results of the primers along with pre-specified cutoff criteria for amplicon product sizes and mismatch criteria to aggregate candidate PCR products from a target database. This pipeline was modified to better accommodate the use of short probe sequences that may contain modifications such as minor-groove-binders by decomposing the problem into a BLAST-based estimate of amplicon products returned by the primers, followed by a Smith-Waterman local alignment of the probe sequences against the predicted amplicon products. Because of the computational cost of alignment with expanding degeneracies, predicted amplicons that contain “N” degenerate characters are not aligned and not considered as inclusive hits. Probe alignments that meet the designated mismatch threshold are annotated to the corresponding amplicon product. This amplicon table is then evaluated using a custom Python script for inclusivity by the criteria above, where set membership for a set of oligos is defined as forward primer, reverse primer and probe sequence are all aligned within the pre-specified mismatch cutoffs. The output from this script is a summary table that gives inclusivity estimates for each unique label, in this case, the Pangolin lineage. Cutoff criteria set inclusion for all assays was a maximum of 3 mismatches in either the forward or reverse primer, 3 mismatches for standard (non-LNA) probes, and a 0 mismatch tolerance for LNA probes.

### MRIGlobal analytical testing

Assay inclusivity/exclusivity testing for all triplexes was performed using serial dilutions of RNA extracted from live SARS-CoV-2 viral stocks, whereas Omicron pentaplex testing was performed using synthetic gRNA from Twist Biosciences due to lack of available stocks for Omicron sublineages. Viral stocks were extracted using the Qiagen Viral RNA Mini Kit (Qiagen, MD, USA). A 10-fold dilution series of each extract was made and then screened for quality/quantity using the Biomeme SARS-CoV-2 Dx assay on a Bio-Rad CFX96 PCR instrument. Dilutions producing Cq values in the early 20s to late 30s (or negative results) were selected for further testing with the variant multiplexes, side-by-side with the Biomeme SARS-CoV-2 Dx assay as a control/comparator assay. This testing was performed using lyophilized versions of Triplexes 2 and 3, but “wet” versions of Triplex 1, Triplex 4, and the Omicron pentaplex were used because final lyophilized versions were not available. “Wet” versions were prepared using commercially available frozen master mix (TaqPath 1-Step RT-qPCR Master Mix, CG from ThermoFisher (St. Louis, MO, USA). When “wet” versions were used, the thermocycling protocol changed to the following: UNG incubation at 25°C for 2 minutes, reverse transcription at 50°C for 15 minutes, initial denaturation at 95°C for 2 minutes, and 45 cycles of 95°C for 3 seconds and annealing/extension at 62°C for 30 seconds. In addition to testing the full dilution series on the Bio-Rad CFX96, single-level testing of each extract was also performed on the handheld Biomeme Franklin™ PCR device. Viral copy numbers for gRNA dilutions were later determined by droplet digital PCR (ddPCR) on a Bio-Rad QX200 (Hercules, CA, USA) and CDC 2019-nCov N1 assay primers and probe.

### MRIGlobal testing of RADx residual clinical specimens

Twenty SARS-CoV-2 residual clinical specimens were provided by the National Institutes of Health (NIH) Rapid Acceleration of Diagnostics (RADx^®^) program. These samples consisted of heat inactivated nasopharyngeal swabs in transport media that had previously tested positive for SARS-CoV-2 and had been sequenced to determine lineage. Five samples each of four different lineages—Alpha, Beta, Gamma, and Delta—were provided by NIH RADx^®^ in early December of 2021 (before Omicron samples were available). Specimens were extracted using the Qiagen Viral RNA Mini kit using 140 µL of sample as input and an elution volume of 70 µL. Extracts were tested with all four variant triplexes, as well as with the Biomeme SARS-CoV-2 Dx assay as a control/comparator assay. This testing was performed on a CFX96 instrument using 2 µL of extract per reaction, and testing duplicate PCR reactions of each sample per triplex. Sample identities were blinded to the operators until after data analysis and interpretation was performed.

### Naval Health Research Center (NHRC) clinical sample testing

Clinical samples tested by NHRC consisted of nasal and nasopharyngeal (NP) swabs in either transport media or saline. For sensitivity and specificity testing, clinical specimens were previously genotyped by whole genome sequencing using the Illumina MiSeq and RNA Prep with Enrichment Library Prep Kit (San Diego, CA, USA). Ten samples were selected for each variant targeted by the panel except for B.1.351, for which NHRC only had four samples. An additional 20 variants not targeted by the panel were also selected to test for specificity of the variant panels. Viral RNA extraction was performed on 100 µl samples using the Qiagen Viral RNA Mini Kit. Extracts were tested with the CDC 2019-nCoV N1 and N2 assays on an ABI 7500 real-time PCR system (Foster City, CA, USA) to confirm presence of SARS-CoV-2 RNA. Variant panel testing was also conducted on the ABI 7500 using TaqPath 1-Step RT-qPCR Master Mix and the variant panel primers and probes provided by MRIGlobal at a 10X concentration. After initial variant panel testing, high-throughput testing was performed for >1500 samples using Triplex 4 (the Omicron triplex). During high-throughput testing, all samples were tested with Triplex 4, and in some cases, only samples that were negative for Triplex 4 assays were then tested with the CDC N1 and N2 assays to verify presence/absence of SARS-CoV-2 in the sample. Many of these samples were later sequenced to confirm the PCR-based lineage determinations.

## RESULTS

### Assay target selection

The Biomeme Franklin™ Real-Time Thermocycler can accommodate three different fluorescence channels, enabling one triplex assay per well. Based on the high prevalence of unique markers among the variant sequences available at the time of assay design, we selected markers to include in a triplex. These markers were either a single nucleotide polymorphism (SNP), multiple SNPs in close proximity, or an insertion or deletion (indel) spanning the probe region. For Triplexes 1 and 2, markers were selected in a way that some are shared among variant lineages and others are unique to a specific lineage to allow for identifying three variants using six markers. For Triplexes 3 and 4, markers were selected for specific identification of a single variant per triplex (Delta and Omicron, respectively). The assay target markers with the algorithm for adjudication of the results and variant calling are presented in Table 1. Data were obtained from outbreak.info (40).

**Table 1.**
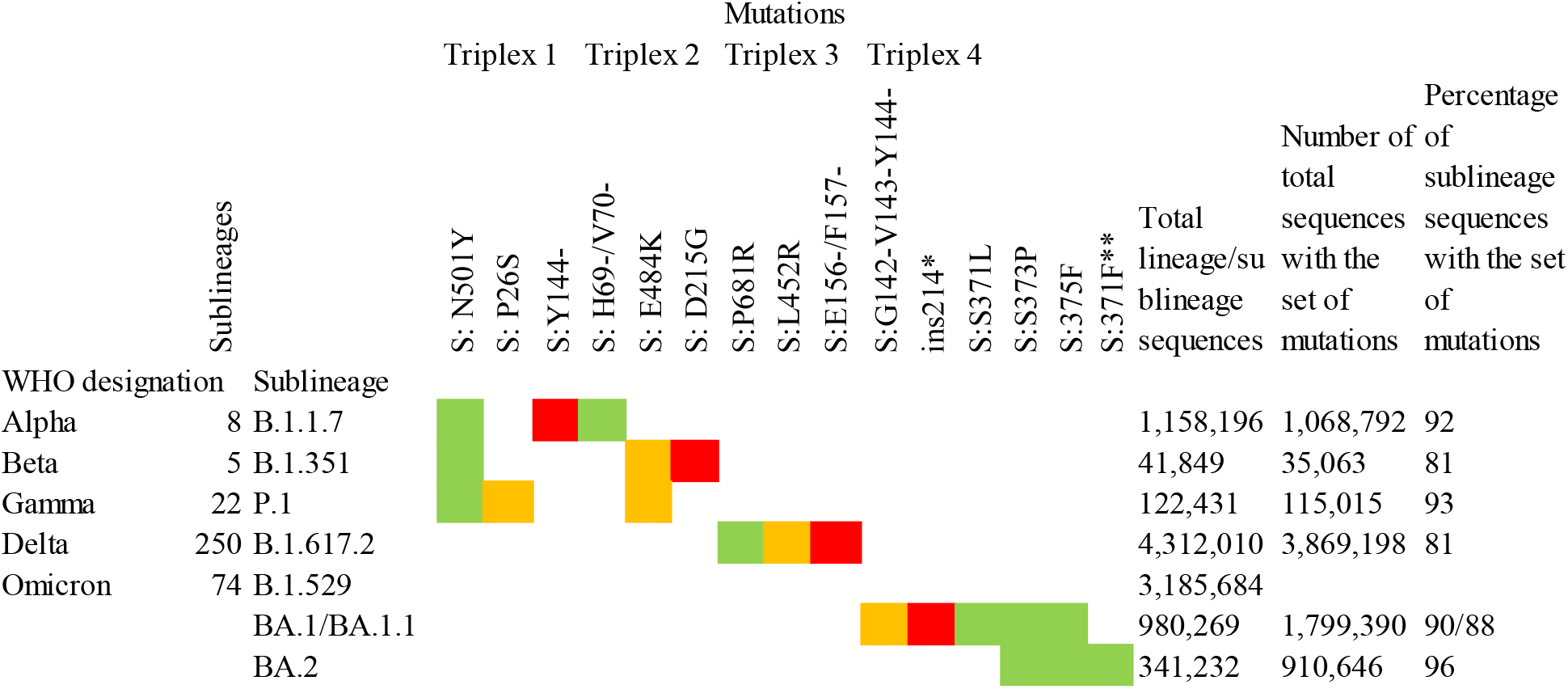
SARS-CoV-2 variant genotyping panel

Total sequences as of April 26,2022: 10,103,411. Source: Outbreak.info Accessed on 04/29/2022 (40). *Outbreak.info can’t currently generate statistics for insertions like the S:ins214EPE mutation. **The S:371-375 signature has a different variation in BA.2 and BA.3 (371F) relative to BA.1 (371L), but is still detected by the same assay despite the single-nucleotide difference. The different colors in the cells represent the corresponding fluorescent channels and probes (Green = FAM, Amber = TEX615, Red = Cy5).

Prevalence and distribution of sequences carrying the triplex specific markers are also shown in the last three columns of Table 1. The total number of sequences of the variant of concern and some of the sublineages and the total number of sequences with a given set of markers are also indicated. The last column represents the percentage of sequences of the specific sublineage listed in column 2 that carries the set of expected mutations. All triplexes cover 81-96% of the specific sublineages they were designed to detect/distinguish. Since the time of assay design, many new sublineages have emerged, including Omicron sublineages BA.3, BA.4, and BA.5. With the increasing number of sublineages and diversity of markers combined with the convergent evolution of markers, the assignment to sublineages based on just two or three markers has become a challenge. In order to address this issue and to increase the confidence of calls for Omicron sublineages, we expanded the panel to a pentaplex assay described in a later section.

### Assay design

The rationale for assay design was to amplify and detect the target amplicon only if the mutation was present in the template. This was accomplished by designing dual-labelled fluorescent probes that can discriminate between the variant markers and the wild-type sequence, in addition to optimizing the annealing temperature and concentration of the probes. For SNP markers in particular, specificity was accomplished using LNA probe designs. As a first step towards assay design, the primer/probe sequences were examined *in silico* to assess the sensitivity and specificity of the designs against whole genome sequences of SARS-CoV-2 variants obtained from GenBank and GISAID (Table 2).

**Table 2.** *In silico* analyses of the assay signature sequences

| Lineage: | B.1.1.7 | B.1.351 | P.1 | B.1.617.2 | BA.1 | BA.1.1 | BA.2 | BA.3 | BA.4 | BA.5 |
| --- | --- | --- | --- | --- | --- | --- | --- | --- | --- | --- |
| Sequences: | 95,621 | 738 | 6,443 | 10,507 | 87,849 | 170,416 | 9,040 | 4,370 | 1,174 | 663 |
| S:N501Y | 98% | 92% | 100% | 0.0% | 92% | 92% | 96% | 99% | 92% | 96% |
| S:P26S | 0.1% | 0.1% | 100% | 0.0% | 0.0% | 0.0% | 0.0% | 0.0% | 0.0% | 0.0% |
| S:Y144del | 98% | 0.4% | 0.2% | 0.0% | 0.0% | 0.0% | 0.1% | 0.1% | 0.0% | 0.0% |
| S:HV69-70del | 98% | 0.0% | 0.0% | 0.2% | 96% | 93% | 0.1% | 68% | 98% | 98% |
| S:E484K | 0.2% | 93% | 97% | 0.0% | 0.0% | 0.0% | 0.0% | 0.0% | 0.0% | 0.0% |
| S:D215G | 0.0% | 98% | 0.0% | 0.0% | 0.0% | 0.0% | 0.0% | 0.0% | 0.0% | 0.0% |
| S:P681R | 0.0% | 0.0% | 0.0% | 99% | 0.0% | 0.0% | 0.0% | 0.6% | 0.0% | 0.0% |
| S:L452R | 0.0% | 0.3% | 0.0% | 95% | 0.2% | 0.0% | 0.1% | 0.6% | 87% | 96% |
| S:EF156-157del | 0.0% | 0.1% | 0.0% | 89% | 0.1% | 0.0% | 0.0% | 0.3% | 0.0% | 0.0% |
| S:S371-375 | 0.0% | 0.0% | 0.0% | 0.0% | 96% | 98% | 96% | 100% | 98% | 98% |
| S:GVY142-144del | 0.0% | 0.0% | 0.0% | 0.1% | 96% | 94% | 0.0% | 100% | 0.0% | 0.0% |
| S:ins214EPE | 0.0% | 0.0% | 0.0% | 0.1% | 79% | 86% | 0.0% | 0.0% | 0.0% | 0.0% |
| N:ERS31-33del | 0.0% | 0.0% | 0.0% | 0.0% | 97% | 97% | 96% | 100% | 91% | 96% |

The *in silico* results indicated very high specificity among the variants of concern, with ≤0.6% of sequences for a particular lineage containing any marker not typically associated with that lineage. Predicted sensitivity was >90% for most of the markers, although there were some exceptions for Delta and Omicron lineages. This reduction in sensitivity is generally a reflection of absence of the mutations (as opposed to PCR failure), as there can be differences among sequences of the same lineage and even among sublineages of the same variant. In some instances, there are shared mutations in some lineages or sublineages due to convergent evolution. For example, S:N501Y was initially designed as a shared mutation for the three variants targeted in Triplexes 1 and 2. It is not present in the Delta sequences, but the majority of Omicron sequences carry this mutation. However, this assay failed to produce detection during analytical testing with Omicron (see Table 3) due to mismatches in the forward and reverse primer regions (Figure S1). These results were further confirmed by clinical sample testing, during which no positive results were obtained for the S:N501Y assay with Omicron samples (see Tables S4 and S5). It is noted that when primers were redesigned to incorporate mixed bases at the mismatch sites, the S:N501Y mutation in Omicron could then be reliably detected (unpublished data.) Similarly, the S:69/70del mutation is present in both Alpha and Omicron BA.1; however, in the latter, there is an additional SNP in the probe binding region (Figures S2 and S3). The S:69/70del probe used in Triplex 2 can still detect Omicron albeit at reduced amplitude, while a more Omicron specific probe can be discriminatory between Alpha and Omicron templates (Figures S4 a-c).

### Analytical testing of Triplexes 1-4

Initial assay testing and optimization was performed using dsDNA templates and/or *in vitro* transcribed RNA templates harboring the targeted regions (data not shown). Once the assays and reaction conditions were finalized, analytical and clinical testing were performed. Analytical testing of the triplex assays was performed using a serial dilution of live viral stocks after extraction as described in the Materials and Methods section. All assays exhibited the expected results for each lineage. Average Cq values (n=2) are presented for dilutions that produced a Cq value of around 22-25 (Table 3). Comparable values were observed between the diagnostic assays (SARS-CoV-2 Dx: spike and Orf1ab) and the variant specific assays.

**Table 3:**
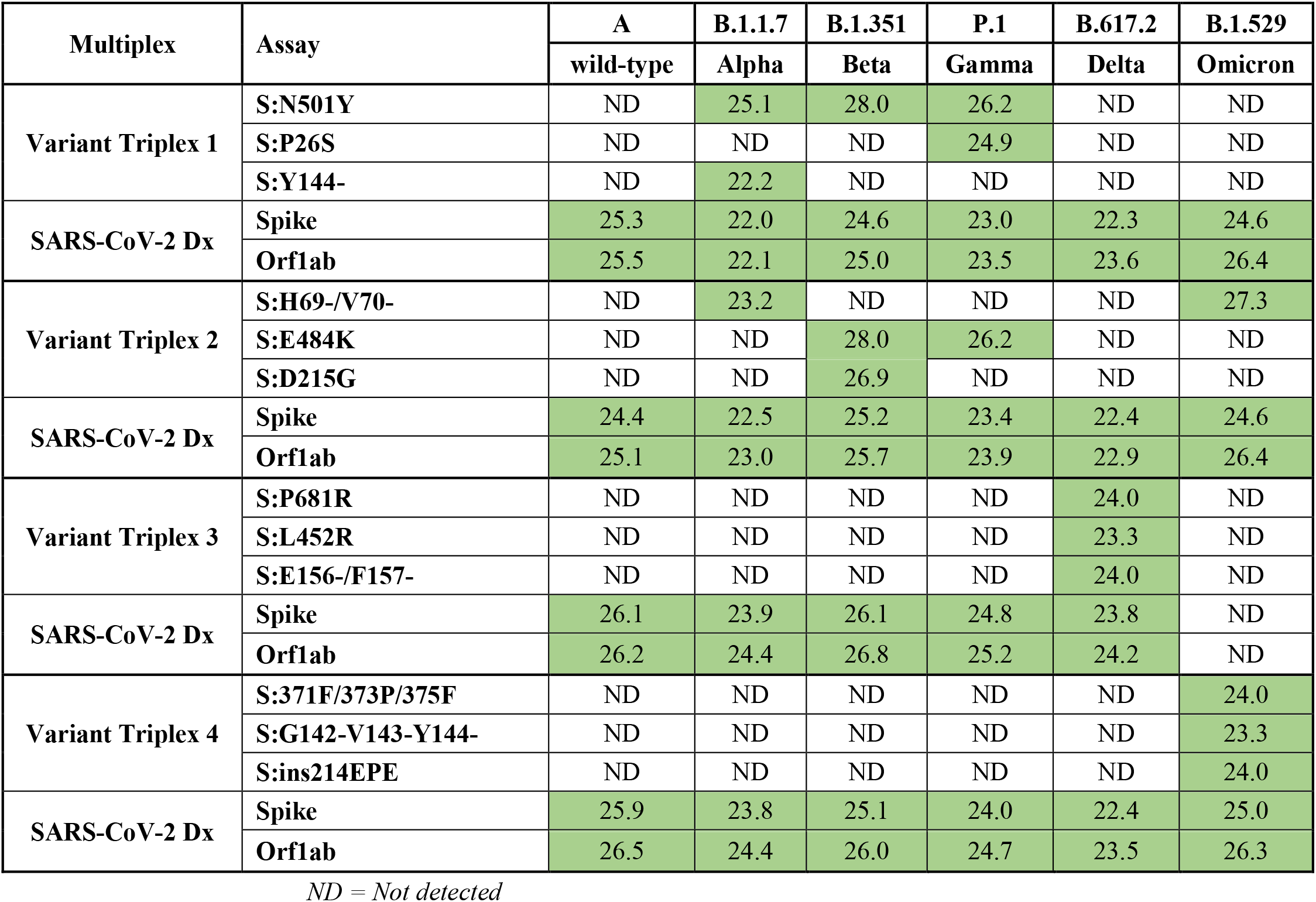
Testing the specificity of Triplexes 1-4 using SARS-CoV-2 viral gRNA

| Multiplex | Assay | A | B.1.1.7 | B.1.351 | P.1 | B.617.2 | B.1.529 |
| --- | --- | --- | --- | --- | --- | --- | --- |
|  |  | wild-type | Alpha | Beta | Gamma | Delta | Omicron |
| Variant Triplex 1 | S:N501Y | ND | 25.1 | 28.0 | 26.2 | ND | ND |
|  | S:P26S | ND | ND | ND | 24.9 | ND | ND |
|  | S:Y144- | ND | 22.2 | ND | ND | ND | ND |
| SARS-CoV-2 Dx | Spike | 25.3 | 22.0 | 24.6 | 23.0 | 22.3 | 24.6 |
|  | Orf1ab | 25.5 | 22.1 | 25.0 | 23.5 | 23.6 | 26.4 |
| Variant Triplex 2 | S:H69-/V70- | ND | 23.2 | ND | ND | ND | 27.3 |
|  | S:E484K | ND | ND | 28.0 | 26.2 | ND | ND |
|  | S:D215G | ND | ND | 26.9 | ND | ND | ND |
| SARS-CoV-2 Dx | Spike | 24.4 | 22.5 | 25.2 | 23.4 | 22.4 | 24.6 |
|  | Orf1ab | 25.1 | 23.0 | 25.7 | 23.9 | 22.9 | 26.4 |
| Variant Triplex 3 | S:P681R | ND | ND | ND | ND | 24.0 | ND |
|  | S:L452R | ND | ND | ND | ND | 23.3 | ND |
|  | S:E156-/F157- | ND | ND | ND | ND | 24.0 | ND |
| SARS-CoV-2 Dx | Spike | 26.1 | 23.9 | 26.1 | 24.8 | 23.8 | ND |
|  | Orf1ab | 26.2 | 24.4 | 26.8 | 25.2 | 24.2 | ND |
| Variant Triplex 4 | S:371F/373P/375F | ND | ND | ND | ND | ND | 24.0 |
|  | S:G142-V143-Y144- | ND | ND | ND | ND | ND | 23.3 |
|  | S:ins214EPE | ND | ND | ND | ND | ND | 24.0 |
| SARS-CoV-2 Dx | Spike | 25.9 | 23.8 | 25.1 | 24.0 | 22.4 | 25.0 |
|  | Orf1ab | 26.5 | 24.4 | 26.0 | 24.7 | 23.5 | 26.3 |

ND = Not detected

For each variant gRNA dilution series and the relevant assays, plots of Cq values versus viral copies per reaction are presented in Figure 2. The PCR efficiencies are presented in Table S6. Amplification efficiencies of all assays with the tested lineages were >80%, with two exceptions (P26S at 79%, and 156/157del at 76%).

**Figure 1.**
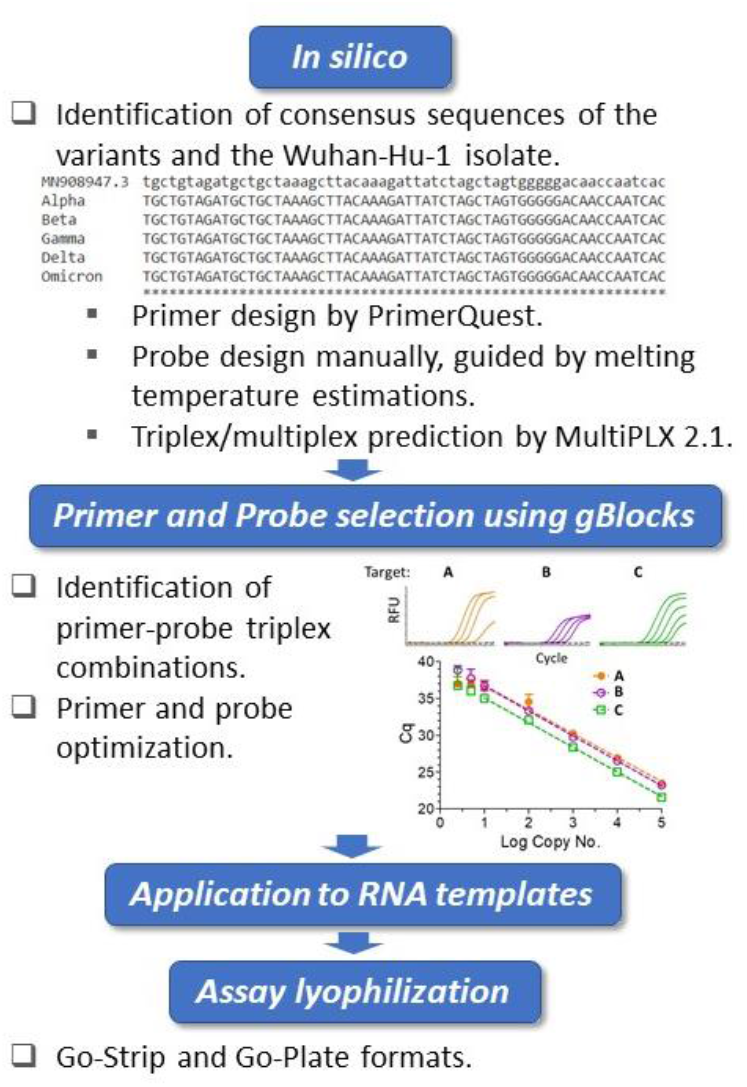
Triplex assay development workflow. Using the Wuhan-Hu-1 isolate (accession no. MN908947.3) as the reference, whole genomes of SARS-CoV-2 variants were compared. The *in silico* framework consisted of identification of appropriate primers and probes and their subsequent suitability in triplex combinations. The primers were designed at regions of consensus sequences, and the probes were designed manually at target mutation sites. For the locked nucleic acid (LNA) probes, melting temperature predictions served as guides for mismatch discrimination. Candidate primers and probes were subsequently aligned using BLAST to ensure specificity, and suitable triplex combinations were subsequently predicted. Identified triplexes were then tested and optimized initially using dsDNA templates (IDT gBlocks), followed by the use of *in vitro*-transcribed RNA templates. The optimized conditions were formulated into freeze-dried Go-Strips and Go-Plates for use on the Biomeme Franklin™ and standard 96-well RT-qPCR instruments, respectively.

**Figure 2:**
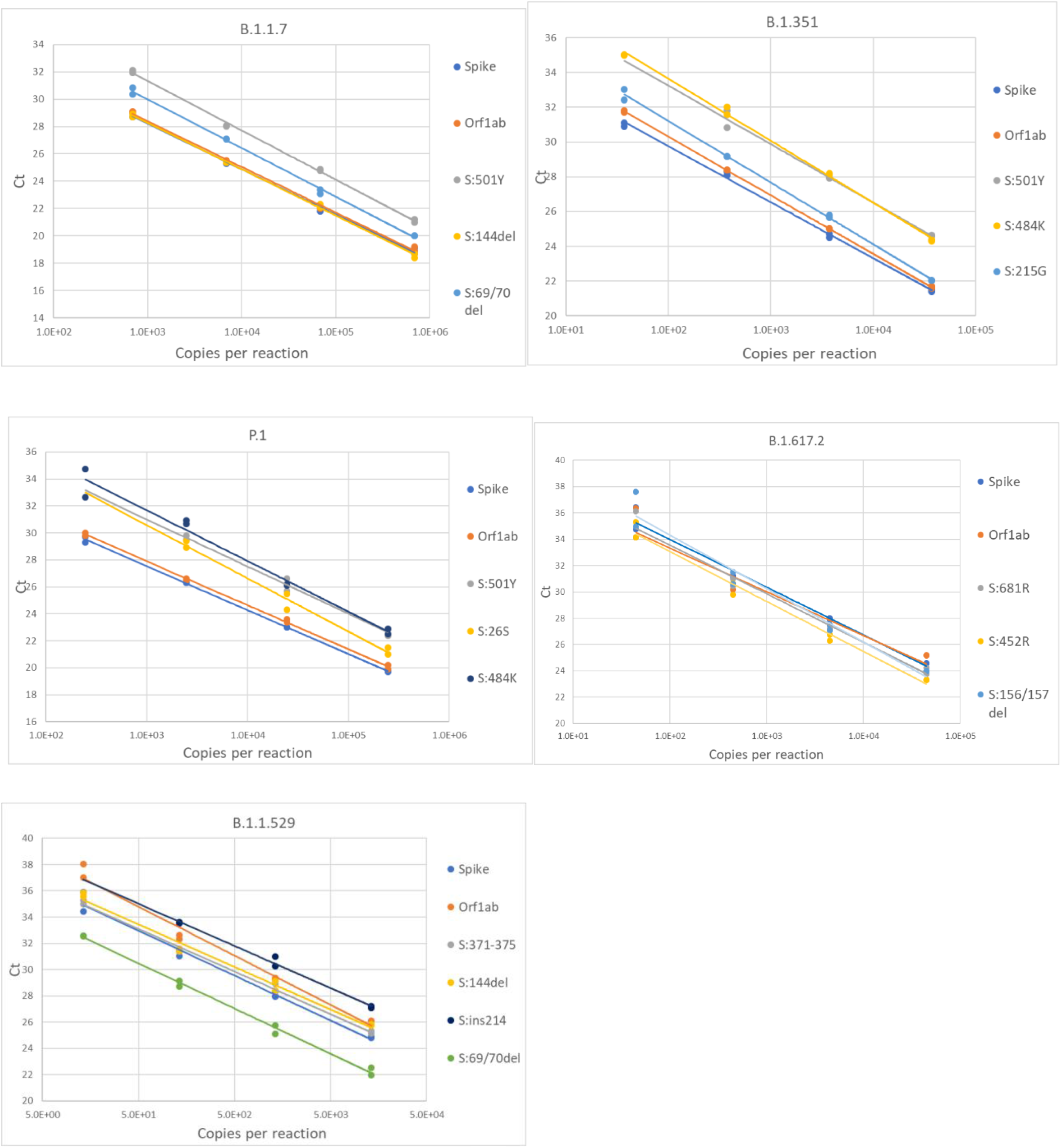
Serial dilution of extracted gRNA of different SARS-CoV-2 variants were used as templates in RT-qPCR reactions. The Cq values are plotted as a function of the concentrations. PCR efficiencies were derived from the slopes of the curves.

### Clinical sample testing

Retrospective testing of the four triplexes using clinical samples was performed as described in the Materials and Methods section. The results are shown in Table 4 and the detailed sample data with Cq values are provided in Tables S4 and S5. The 64 samples presented in Table 4 are from two different sources: NIH RADx^®^ and NHRC. Overall, the panels have very high sensitivity and specificity (see Figures S5 a,b). However, in this analysis results of the S:P26S assay in Triplex 1 have been excluded due to indiscriminate signal (and hence false positives) with all SARS-CoV-2 samples analyzed on the ABI 7500 (See Figures S5-b). Therefore, positive calls for the P.1 lineage are based on only two markers instead of three.

**Table 4.** Sensitivity and specificity of Triplexes 1-4 based on clinical sample testing

| Lineage | Samples in the test set | True positives | False positives | Inconclusive (low titer) | Sensitivity | Specificity |
| --- | --- | --- | --- | --- | --- | --- |
| B.1.1.7 | 15 | 14 | 0 | 1 | 93.3% | 100.0% |
| B.1.351 | 9 | 9 | 0 | 0 | 100.0% | 100.0% |
| P.1 | 15 | 14 | 1 | 1 | 93.3% | 93.3% |
| B.1.617.2 | 15 | 15 | 0 | 0 | 100.0% | 100.0% |
| BA.1 | 10 | 10 | 0 | 0 | 100.0% | 100.0% |
| **OVERALL** | **64** | **62** | **1** | **2** | **96.9%** | **98.4%** |

Samples tested with lineages not targeted by the panel: 20, with only one producing a false positive. P.1 false positive was for a B.1.621 specimen on the basis of having the same two mutations as P.1 when the P26S assay is ignored.

In addition to the 64 samples from targeted lineages, 20 samples (1 per lineage) from lineages not targeted by the panel were also tested, with only one producing a false positive lineage call; a B.1.621 specimen was falsely identified as P.1 on the basis of having the same two mutations as P.1 when the P26S assay is ignored. This was the only false positive call obtained from the test set of 84 clinical specimens used to assess sensitivity and specificity of Triplexes 1-4. Difficulty distinguishing amplification from background for the S:P26S assay was seen both in the NHRC clinical sample testing on ABI 7500 systems (Figures S5-b) and in testing on the Biomeme Franklin™ (Figure 4), but was less of an issue during analytical testing and testing of the RADx clinical specimens on the Bio-Rad CFX96.

### Additional clinical sample testing with Triplex 4

Due to the surge in US clinical samples and testing needs for Omicron, beginning in December of 2021, additional testing was performed on the Omicron triplex (Triplex 4). This consisted of dynamic range finding and relative limit of detection (LoD) testing with a clinical specimen, additional sensitivity and specificity testing with an expanded set of samples, and high-throughput testing of >1500 clinical samples being screened for sequencing.

### Dynamic Range and Relative Limit of Detection (LoD) of Triplex 4

In order to assess the dynamic range and limit of detection of the Triplex 4 panel of assays, a 10-fold serial dilution of a clinical specimen extract was made and PCR was performed as described in the Materials and Methods section. This specimen was known to be SARS-CoV-2 positive and of sublineage BA.1 based on prior PCR and sequencing. Triplicate reactions of Triplex 4 assays were tested for each dilution and the working LoD was determined as the lowest sample level producing 3 of 3 positive results. Working LoD was the same for all three assays in the triplex, with Cq values ranging from 34-37 at that level. This working LoD was 1:1e5 dilution of the specimen extract, which produced N1 and N2 Cq values of ∼18 at the undiluted level.

### Sensitivity and Specificity of Triplex 4

We further assessed sensitivity and specificity of the Triplex 4 using newly obtained clinical samples. Twenty of 20 Omicron (BA.1) clinical specimens tested positive for all three Omicron assays, including three specimens with “failed” sequencing results due to lower titers. The Cq values of CDC N1 and CDC N2 assays ranged from ∼16-30 for these samples. Fourteen of 14 non-Omicron specimens (all of different lineages; see Table S5 for lineages) were negative for all three Omicron assays and positive for the CDC N1 and N2 assays, with Cq values of 22-30. Based on these data for sequence-confirmed samples, the sensitivity and specificity were 100% for all three assays in the panel. Ten samples without sequence data were also tested with Triplex 4. Of these ten unknown samples, eight were positive for all three assays, and two were negative. Of the two negative samples, one was positive for the CDC N1 and N2 assays (suggesting it contained a non-Omicron SARS-CoV-2 lineage) and the other one was negative for the CDC assays. Cq data for all samples are summarized in Figure 3 and Table S7.

**Figure 3:**
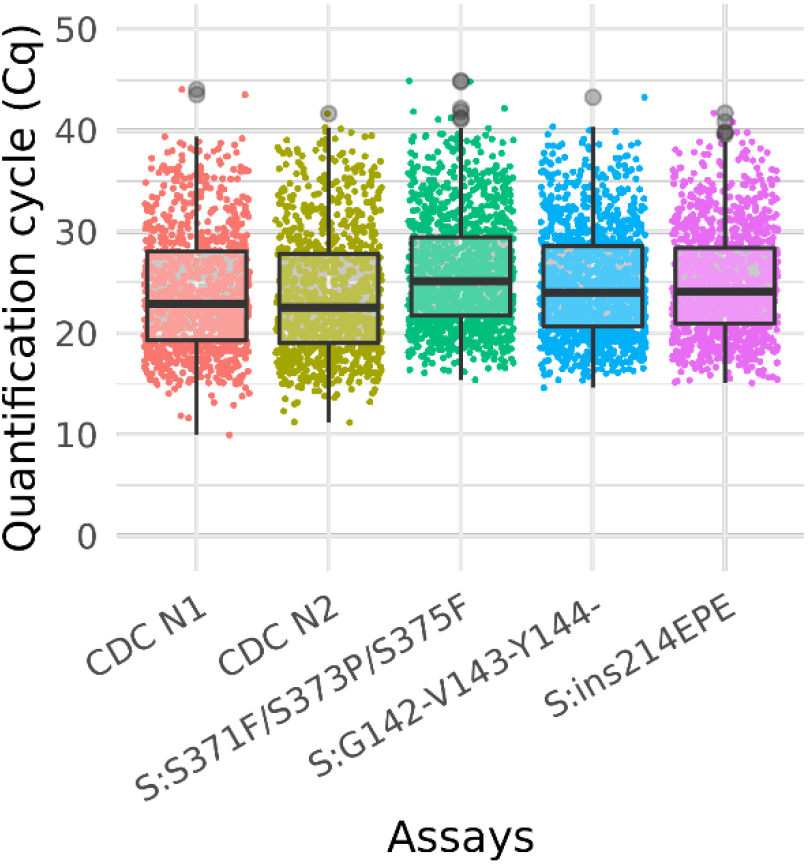
Box plot of Ct values of omicron sample testing results Cq values for clinical samples tested with Triplex 4 and the CDC N1 and N2 assays. The total number of samples is 1548. The positive sample counts for each assay are: CDC N1 (1013), CDC N2 (1014), S:371-375 (1255), S:142-144del (1213), and S:ins214 (1189). Grey circles represent outliers. Note: Not all samples were tested with the CDC assays.

### High-throughput clinical sample testing with Triplex 4

A total of 1548 clinical samples were tested with Triplex 4. Of these, 1255 (81%) were positive for one or more of the Omicron triplex assays. The sample and Cq values of these samples are presented in Table S8 and the results are shown in the box plot (Figure 3).

### Performance assessment of all triplexes on the Biomeme Franklin

Synthetic gRNA obtained from Twist Biosciences for six lineages (B.1.1.7, B.1.351, P.1, B.1.617.2, BA.1, and BA.2) were used to test performance of the variant triplexes on Biomeme Franklin™ devices. The Biomeme Franklin™ is a handheld, battery-powered nucleic acid testing device with 9 wells and 3-color detection that is designed for field portability in low-resource, far-forward environments (36). Negative controls were also included for each assay/run and consisted of water-only NTC reactions, as well as synthetic gRNA from the wild-type (A) lineage. We initially tested each lineage at a concentration of 500 genome equivalent copies (GEC) per reaction. At this level, all assays produced easily discernable amplification for all expected mutations in each lineage, with two exceptions: the S:P26S and S:N501Y assays in which Triplex 1 yielded only dim amplification curves that were not consistently detected (Figure 4a). When Triplex 1 was retested at a 100-fold higher concentration (50,000 GEC per reaction), these assays did produce positive results, but amplification curves remained dim (Figure 4b). These results are consistent with other results obtained on the Bio-Rad CFX96 and ABI 7500, with the S:P26S and S:N501Y assays producing dimmer curves and demonstrating less sensitive detection (higher LoDs) than the other assays in the panel, nonetheless detectable. All NTC reactions and other expected negatives were negative across all runs.

**Figure 4.**
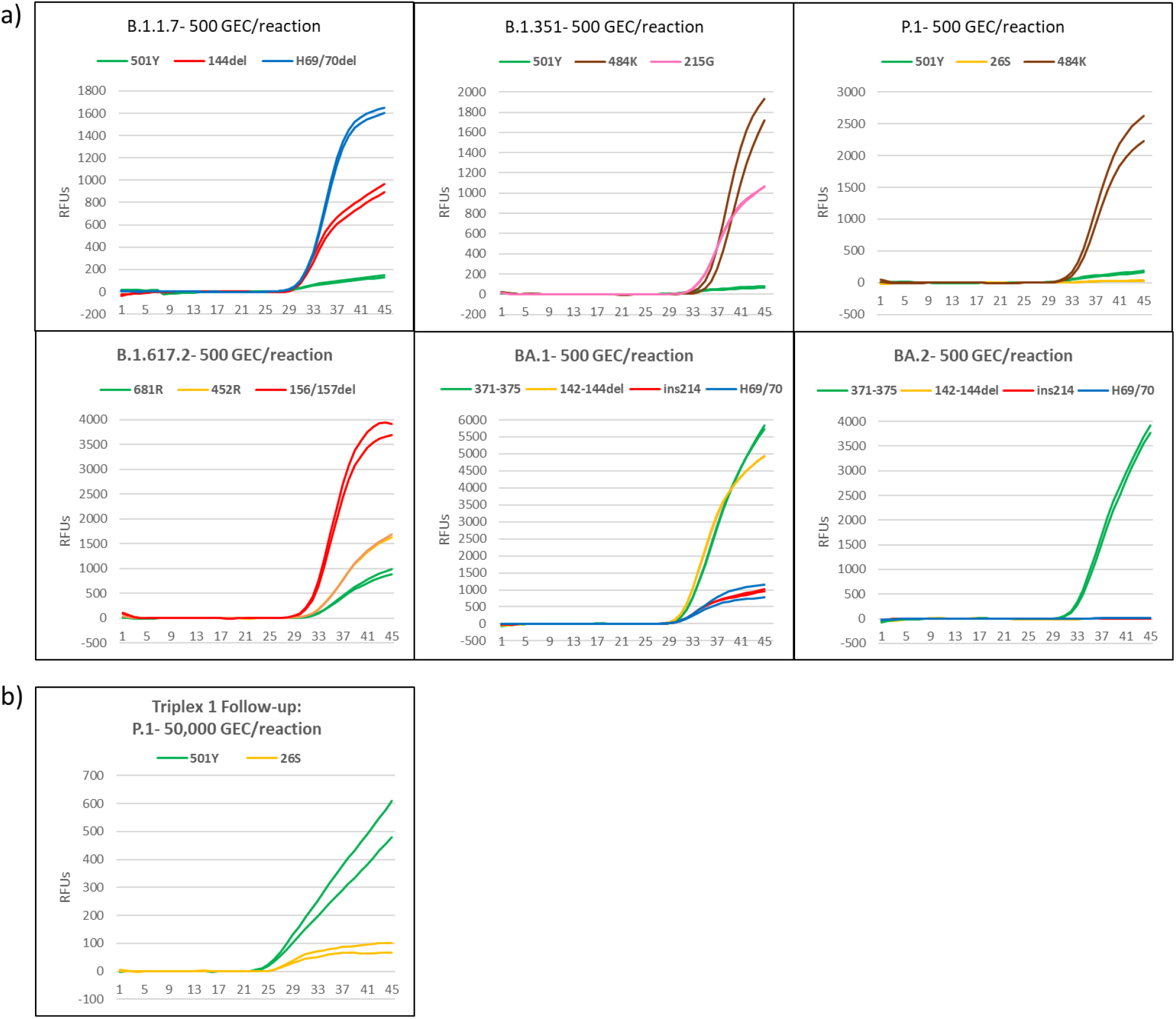
Amplification plots of Triplexes 1-4 on the Biomeme Franklin™ device. A) Plots of templates with 500 genomic copy equivalents of different variants. B) Amplicfication plot of P1 template at 50,000 copies.

### Omicron Pentaplex Analytical Testing

Omicron pentaplex testing was performed using serial dilutions (50 to 50,000 copies per reaction) of gRNA for BA.1 and BA.2, and of gBlocks for BA.1.1, BA.3, BA.4, and BA.5 due to unavailability of gRNA for those lineages. Wild-type gRNA was used as a negative control. For comparison, these serial dilutions were also tested with Triplexes 1-4. All results were as expected based on *in silico* predictions and known variant mutation profiles according to outbreak.info (40). For positive samples, average Cq values (n=2) are provided for the 50 copies per reaction test level (Table 6). Use of either the Omicron pentaplex or a combination of Triplexes 2-4 allowed for discrimination among BA.1/BA.1.1, BA.2, BA.3, and BA.4/BA.5, whereas use of only the original Omicron triplex (Triplex 4) would not have allowed for discrimination of BA.2 from BA.4 and BA.5. The BA.4 and BA.5 sublineages (as well as some sublineages of BA.2) also have an additional marker of clinical significance, S:L452R, which is part of Triplex 3 and can be used for further discriminatory power (41).

**Table 5.** Dynamic range testing

|  | S:S371L, S373P, & S375F |  |  | S:del142-144 |  |  | S:ins214EPE |  |  |
| --- | --- | --- | --- | --- | --- | --- | --- | --- | --- |
|  | Rep 1 | Rep 2 | Rep 3 | Rep 1 | Rep 2 | Rep 3 | Rep 1 | Rep 2 | Rep 3 |
| **Undiluted** | 21.1 | 21.0 | 21.3 | 19.1 | 19.0 | 19.1 | 18.5 | 18.4 | 18.6 |
| **10<sup>-1</sup>** | 24.1 | 24.0 | 23.9 | 22.3 | 22.3 | 22.2 | 21.9 | 21.8 | 21.6 |
| **10<sup>-2</sup>** | 27.3 | 27.4 | 27.1 | 25.5 | 25.5 | 25.4 | 25.0 | 25.1 | 25.0 |
| **10<sup>-3</sup>** | 30.6 | 30.4 | 30.6 | 28.9 | 28.5 | 28.7 | 28.3 | 28.2 | 28.3 |
| **10<sup>-4</sup>** | 33.5 | 33.9 | 34.1 | 32.1 | 32.2 | 32.3 | 31.6 | 31.7 | 31.8 |
| **10<sup>-5</sup>** | **36.8** | **37.4** | **37.2** | **34.7** | **35.6** | **34.3** | **34.6** | **34.6** | **34.2** |
| **10<sup>-6</sup>** | ND | 38.5 | ND | 37.7 | 37.4 | ND | 37.5 | 37.1 | ND |
| **10<sup>-7</sup>** | ND | ND | ND | ND | ND | ND | ND | ND | ND |

ND = Not detected

**Table 6.** Inclusivity/Exclusivity Testing with an Expanded Omicron Panel

| Multiplex | Assay | A<br>(wild-type) | BA.1<br>gRNA | BA.2<br>gRNA | BA.1.1<br>gBlock | BA.3<br>gBlock | BA.4<br>gBlock | BA.5<br>gBlock |
| --- | --- | --- | --- | --- | --- | --- | --- | --- |
| Triplex 1 | S:N501Y | ND | ND | ND | ND | ND | ND | ND |
|  | S:P26S | ND | ND | ND | ND | ND | ND | ND |
|  | S:Y144- | ND | ND | ND | ND | ND | ND | ND |
| Triplex 2 | S:H69-/V70- | ND | 33.0 | ND | 34.4 | 33.5 | 32.5 | 33.5 |
|  | S:E484K | ND | ND | ND | ND | ND | ND | ND |
|  | S:D215G | ND | ND | ND | ND | ND | ND | ND |
| Triplex 3 | S:P681R | ND | ND | ND | ND | ND | ND | ND |
|  | S:L452R | ND | ND | ND | ND | ND | 33.2 | 32.8 |
|  | S:E156-/F157- | ND | ND | ND | ND | ND | ND | ND |
| Triplex 4 | S:371F/373P/375F | ND | 32.0 | 33.6 | 34.0 | 32.9 | 32.4 | 33.3 |
|  | S:G142-V143-Y144- | ND | 32.3 | ND | 33.9 | 33.5 | ND | ND |
|  | **S:ins214EPE** | ND | 33.7 | ND | 34.4 | ND | ND | ND |
| **Omicron Pentaplex** | **S:371F/373P/375F** | ND | 35.6 | 35.0 | 35.8 | 34.9 | 34.3 | 35.0 |
|  | **S:G142-V143-Y144-** | ND | 34.5 | ND | 34.4 | 34.3 | ND | ND |
|  | **S:ins214EPE** | ND | 34.7 | ND | 33.5 | ND | ND | ND |
|  | **S:H69-/V70-** | ND | 34.8 | ND | 36.0 | 34.9 | 36.1 | 36.3 |
|  | **N:E31-R32-S33-** | ND | 35.2 | 35.9 | 34.1 | 34.0 | 34.4 | 34.6 |

ND = Not detected

## DISCUSSION

The emergence of variants with phenotypic attributes may have impacts on the effectiveness of vaccines, therapeutics and diagnostics. The resulting mutant proteins can lead to diminished molecular recognition for vaccine and therapeutic interventions. In regard to diagnostics, signature erosion is a term used for defining the mutational changes in molecular assay signatures such as primer/probe binding sites (42, 43). The term ‘signature erosion’ is used here to signify potential false positive or false negative results in molecular assays due to mutations in the PCR primers/probe/amplicon target sequences (PCR signatures).

In this study, we describe a panel of multiplex PCR assays for use in a point-of-care PCR device as well as other standard PCR platforms targeting a set of mutations across the major SARS-CoV-2 variants of concern; viz., Alpha, Beta, Gamma, Delta and Omicron. The Omicron triplex (Triplex 4) can distinguish among Omicron sublineages BA.1, BA.2, and BA.3, and the Omicron pentaplex can further delineate BA.4 and BA.5. We determined a sensitivity and specificity of >90% for these panels with sequence-confirmed SARS-CoV-2 clinical specimens. Based on these data, we posit that these panels could be valuable tools to fill variant testing needs and serve as a screening test to inform selection/prioritization of samples for whole genome sequencing. In the current configuration, the panels are used as non-Dx assays because of the cost involved in Dx development and are meant to be used as Research Use Only (RUO) assays. The general strategy is to test the samples using the Biomeme EUA Dx assay first followed by screening for variants using the RUO genotyping panel assays. As of May 27, 2022, there are no EUA SARS-CoV-2 genotyping assays and there are several with a sequencing component as part of the assay (4).

There are several distinct advantages to using SARS-CoV-2 variant genotyping assays as a screening tool. 1) PCR is extremely less complex, cheaper and faster compared to sequencing, meaning a PCR panel could be a better alternative for rapid identification and tracking of variants. 2) A genotyping PCR panel could be very useful in resource-limited settings, since sequencing is not done at the same level in all locations/countries, and many laboratories do not have the infrastructure or expertise to perform sequencing. 3) Obtaining whole genome sequence data from clinical samples is very much dependent on viral load; reported PCR Cq cutoffs for obtaining full-length genome sequences range from approximately 25-33 (44, 45). A PCR panel could fill this gap by providing genotype information for numerous samples that cannot be sequenced due to low viral load as reflected by high Cq values. 4) A variant PCR panel can be used as an efficient and cost-effective screening assay to prioritize samples for sequencing, conserve resources, and minimize over/under-sampling of certain lineages for sequencing. 5) Mutation-specific PCR amplification can be a more precise option than a proxy based on assay failure, such as the well-known S gene target failure (SGTF). Unlike SGTF, in which negative results are used as a proxy to indicate presence of variants like Alpha and Omicron BA.1, this variant PCR panel comprises specific assays that amplify only the mutant template and not the wild type. So variant detection is based on a positive PCR amplification of the presence of a variant in the sample. 6) During the course of the pandemic, multiple ‘discrete’ waves of variants emerged; hence, unique variant panels can be very useful to detect variants with no ambiguity when there are multiple variants in the community; the same is true for mixed infections. 7)

Wastewater testing is a good predictor of the impending community prevalence; in fact, a few days to a week in advance of community prevalence (46). But, sequencing wastewater samples to monitor the spread of genotypes is problematic/difficult due to sample quality and the presence of many different strains/genotypes in the same sample, potentially at low-titer, so a PCR panel that can detect variant markers would be ideal for wastewater monitoring. Similarly, the variant panel can also be a useful, cheaper alternative for other types of environmental testing; e.g., contamination/decontamination verification, and other types of non-clinical uses, as RUO assays are typically less expensive than Dx assays. These variant panels can help fulfill the need for rapid identification of the SARS-CoV-2 variant present in a sample, leading to quick decision making with respect to public health measures as well as treatment options for individuals.

## Data Availability

All data produced in the present work are contained in the manuscript and supplemental files. Any additional data are available upon reasonable request to the authors.

## ACKNOWLEDGEMENTS

We would like to acknowledge Eric Lai and the NIH RADx program (https://www.nih.gov/research-training/medical-research-initiatives/radx) for providing some of the clinical samples used in this study. We would also like to thank Robert Player for help generating the Figure 3 box plot.

## Author Contributions

Conceptualization: JvW, JS, MN, SS; Experimental design: JvW, MN, EW, JS, SS; Methodology & Experimental validation: BJS, MN, BP, BK, PD, RAD, RP, AF; Formal analysis: BJS, MN, BP, BK, PD, JS; Data curation & visualization: JS, BP; Resources: CM, JvW, JS; Project Administration & Supervision: JvW, JS, SS; Funding Acquisition: SS, BG; Writing – Original Draft Preparation: SS, JS, MN; Writing – Review & Editing-All authors

## DISCLAIMERS

The content of this publication are the authors’ opinions and do not necessarily reflect the views or policies of the U.S. Department of Defense, the U.S. Department of the Army, or the institutions and companies affiliated with the authors, nor does the mention of trade names, commercial products, or organizations imply endorsement by the U.S. Government. The author reports no potential conflicts of interest.

Some authors of this work are military service members or federal/contracted employees of the United States government. This work was prepared as part of their official duties. Title 17 U.S.C. 105 provides that copyright protection under this title is not available for any work of the United States Government. Title 17 U.S.C. 101 defines a U.S. Government work as work prepared by a military service member or employee of the U.S. Government as part of that person’s official duties.

## APPENDICES

Supplementary Tables

Supplementary Figures

