## Supplemental Figures for "Development, testing and validation of a SARS-CoV-2 multiplex panel for detection of the five major variants of concern on a portable PCR platform"

### Slide 1
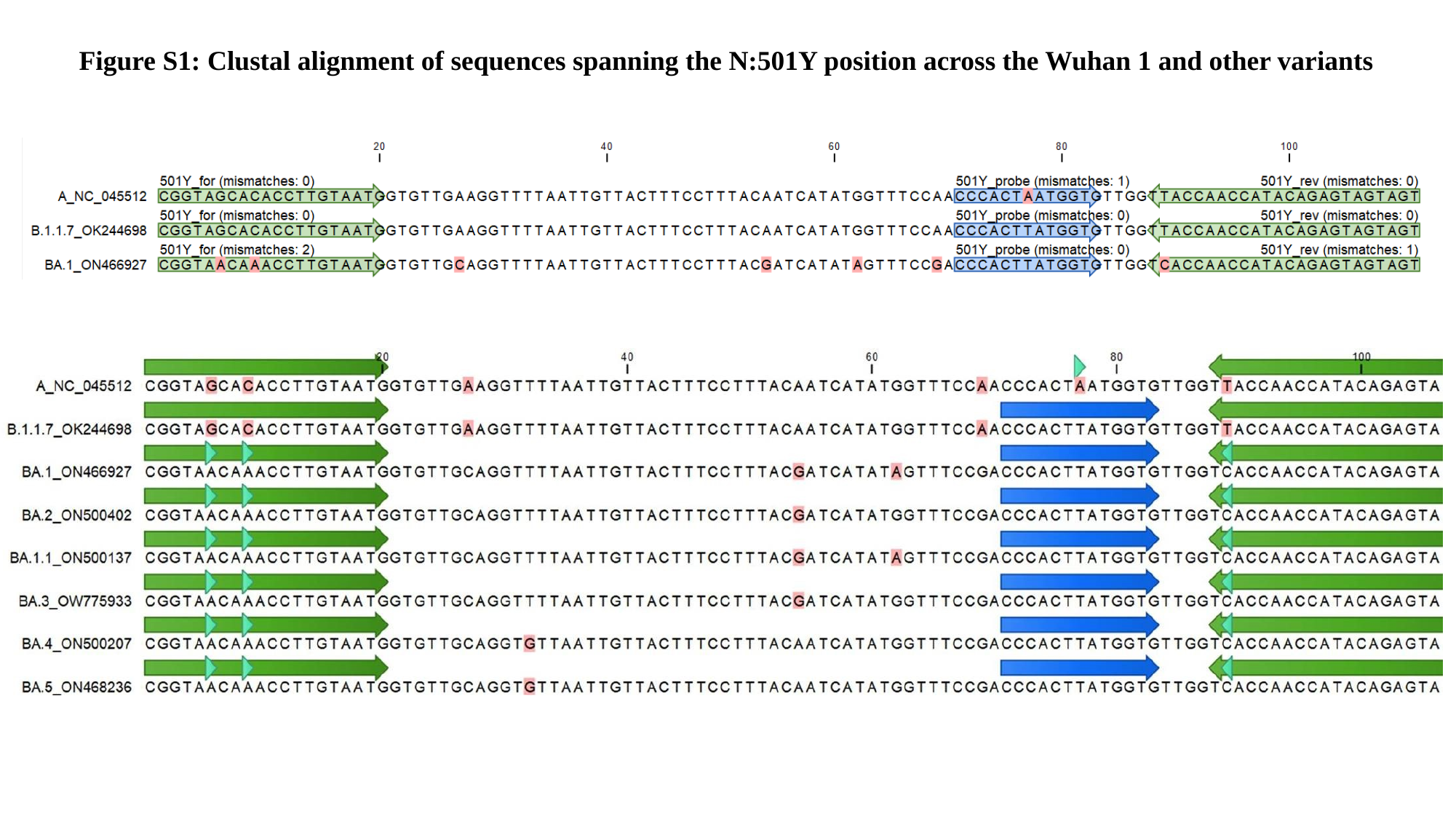

Figure S1: Clustal alignment of sequences spanning the N:501Y position across the Wuhan 1 and other variants

### Slide 2
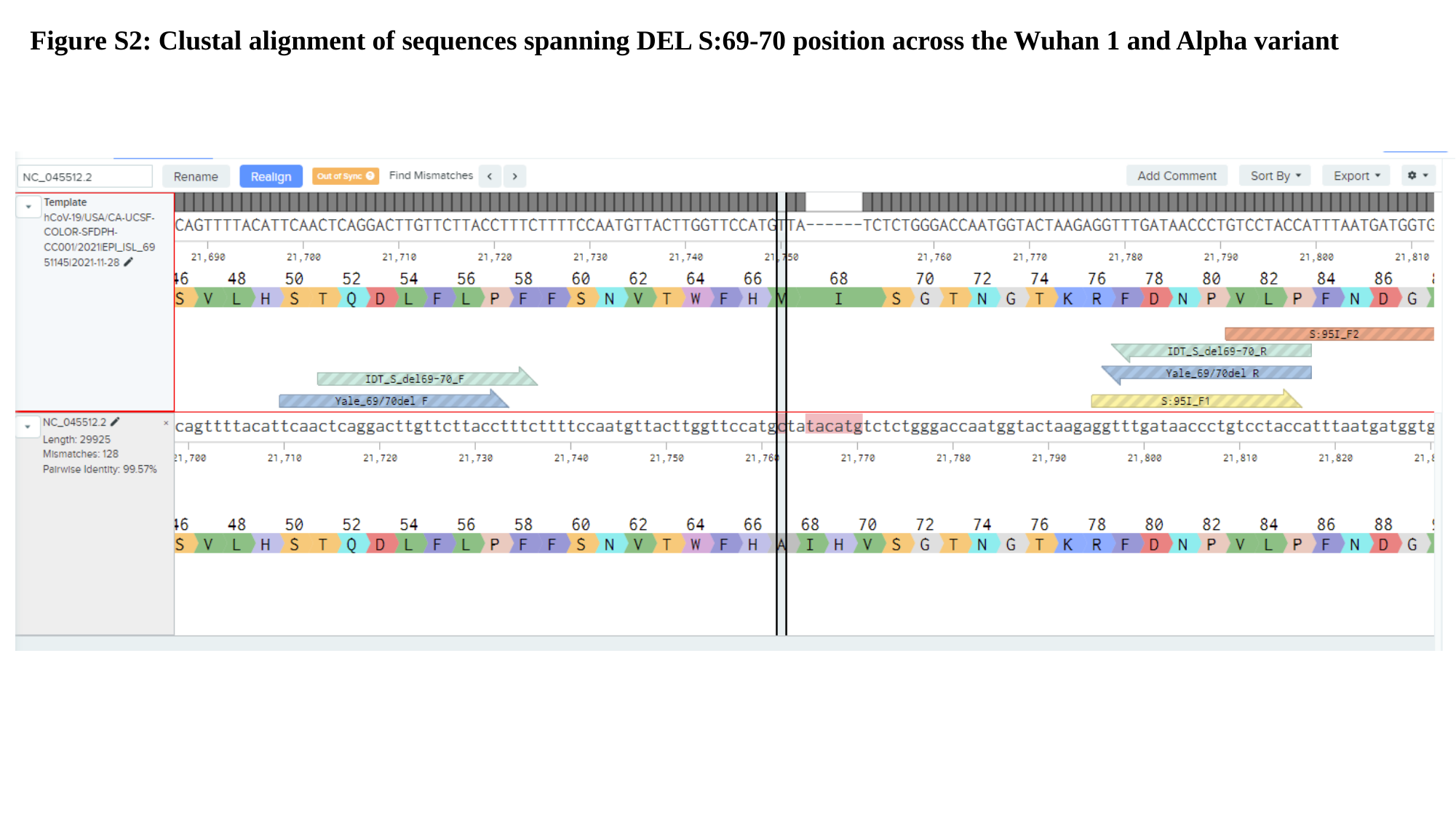

Figure S2: Clustal alignment of sequences spanning DEL S:69-70 position across the Wuhan 1 and Alpha variant

### Slide 3
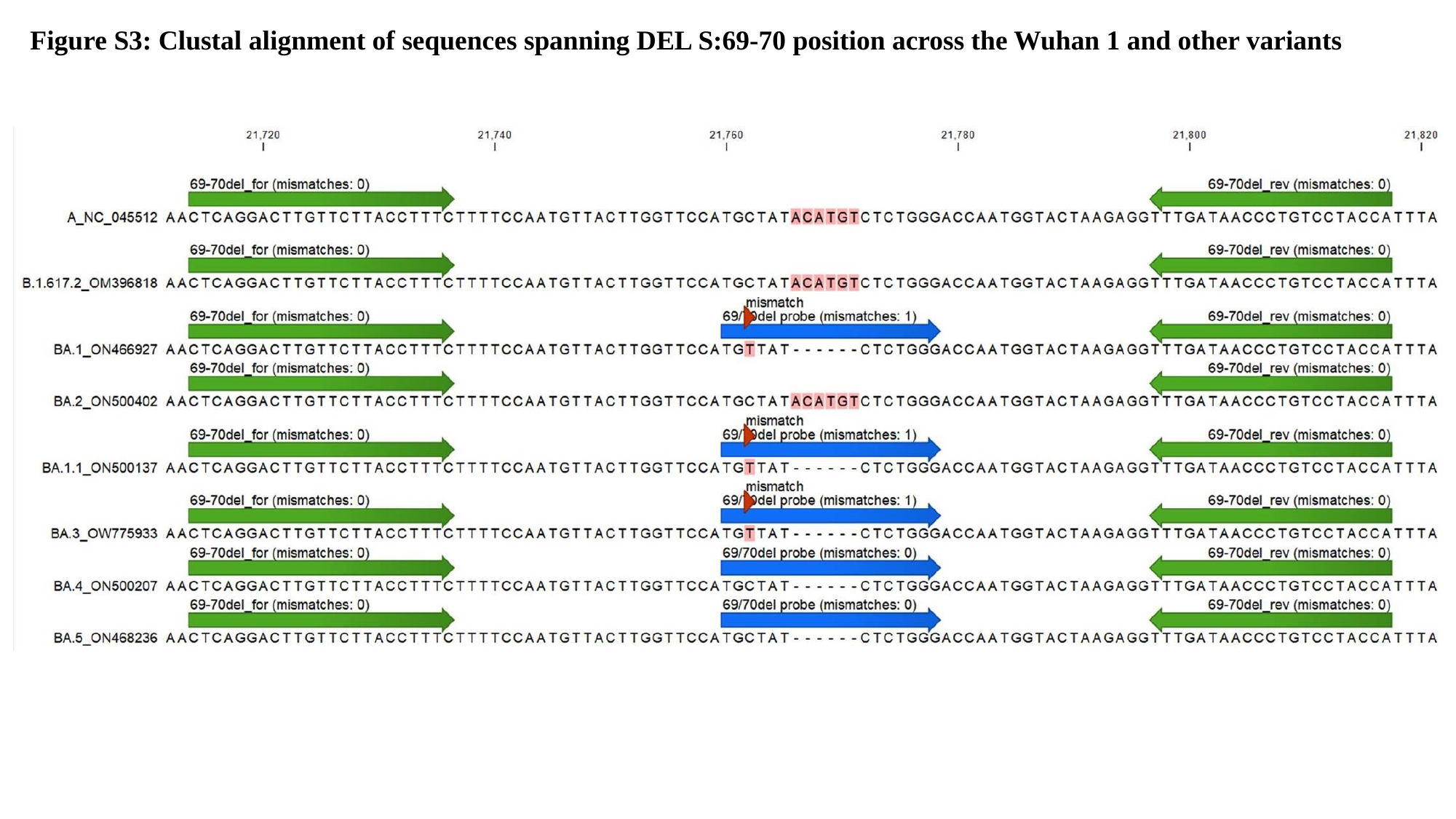

Figure S3: Clustal alignment of sequences spanning DEL S:69-70 position across the Wuhan 1 and other variants

### Slide 4
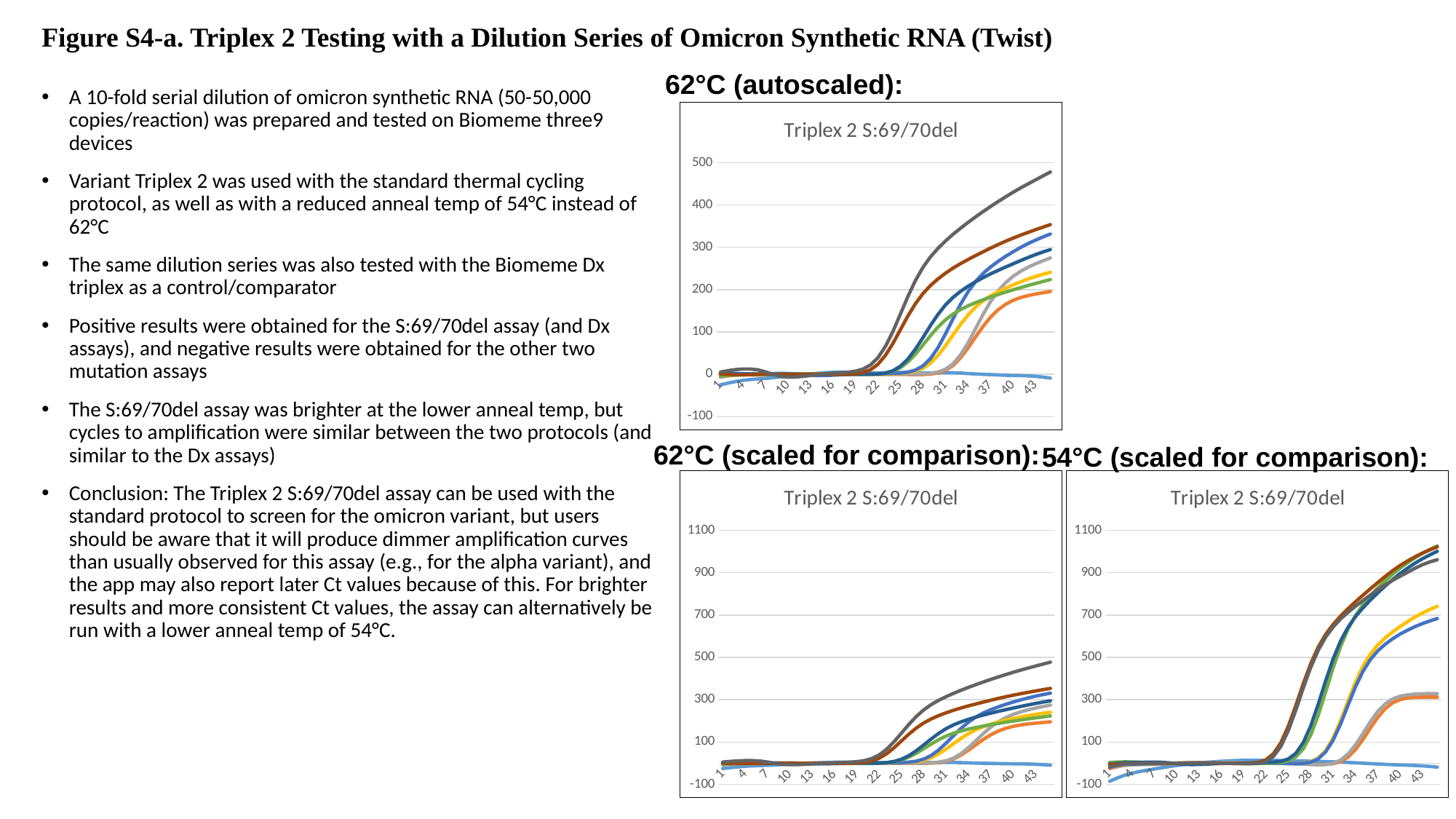

#### Figure S4-a. Triplex 2 Testing with a Dilution Series of Omicron Synthetic RNA (Twist)
62°C (autoscaled):
A 10-fold serial dilution of omicron synthetic RNA (50-50,000 copies/reaction) was prepared and tested on Biomeme three9 devices
Variant Triplex 2 was used with the standard thermal cycling protocol, as well as with a reduced anneal temp of 54°C instead of 62°C
The same dilution series was also tested with the Biomeme Dx triplex as a control/comparator
Positive results were obtained for the S:69/70del assay (and Dx assays), and negative results were obtained for the other two mutation assays
The S:69/70del assay was brighter at the lower anneal temp, but cycles to amplification were similar between the two protocols (and similar to the Dx assays)
Conclusion: The Triplex 2 S:69/70del assay can be used with the standard protocol to screen for the omicron variant, but users should be aware that it will produce dimmer amplification curves than usually observed for this assay (e.g., for the alpha variant), and the app may also report later Ct values because of this. For brighter results and more consistent Ct values, the assay can alternatively be run with a lower anneal temp of 54°C.
###### Chart: Triplex 2 S:69/70del
| Category | NTC | 50 copies | 50 copies | 500 copies | 500 copies | 5,000 copies | 5,000 copies | 50,000 copies | 50,000 copies |
|---|---|---|---|---|---|---|---|---|---|62°C (scaled for comparison):
54°C (scaled for comparison):
###### Chart: Triplex 2 S:69/70del
| Category | NTC | 50 copies | 50 copies | 500 copies | 500 copies | 5,000 copies | 5,000 copies | 50,000 copies | 50,000 copies |
|---|---|---|---|---|---|---|---|---|---|
###### Chart: Triplex 2 S:69/70del
| Category | Well 1 green | Well 2 green | Well 3 green | Well 4 green | Well 5 green | Well 6 green | Well 7 green | Well 8 green | Well 9 green |
|---|---|---|---|---|---|---|---|---|---|

### Slide 5
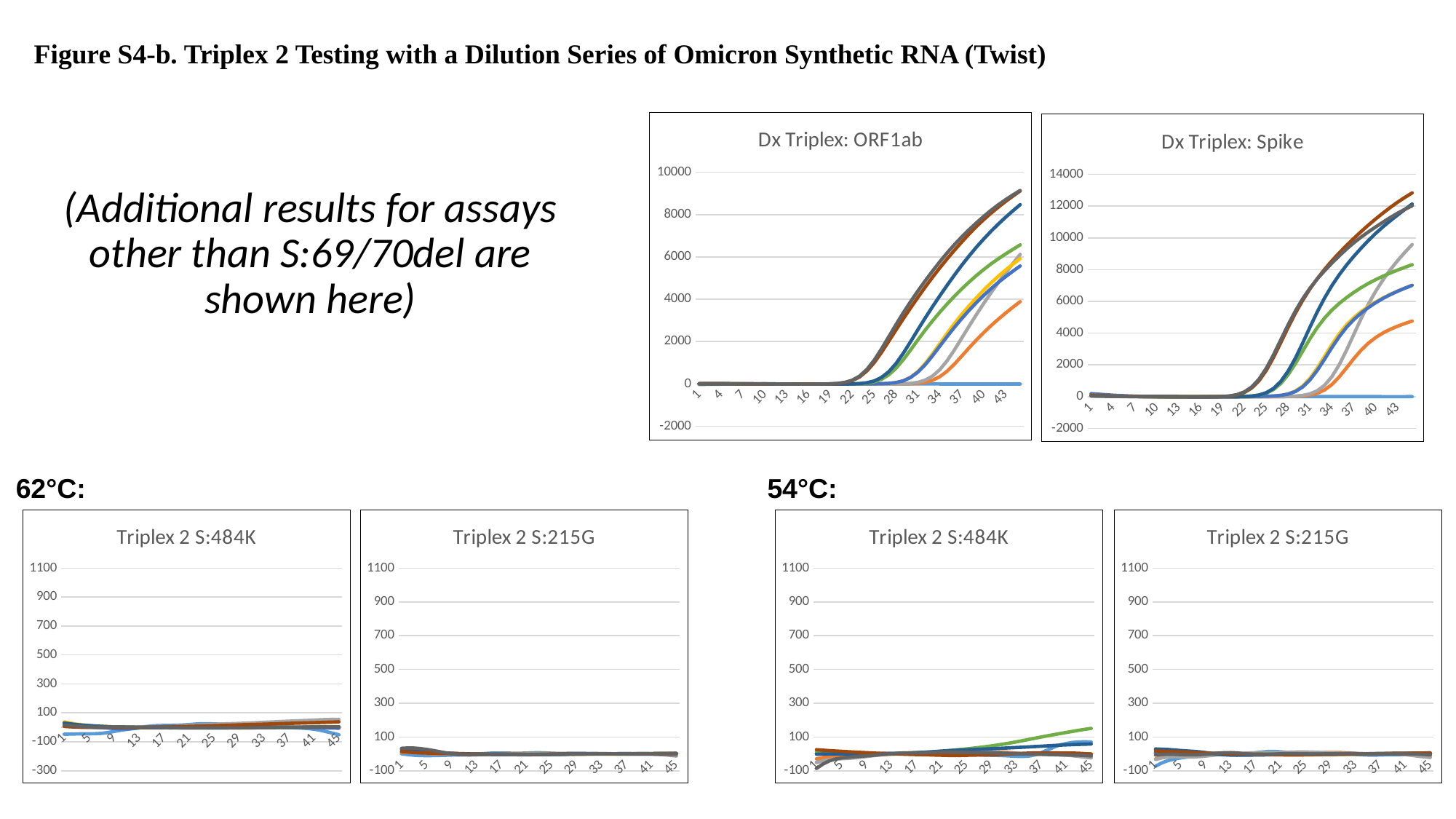

#### Figure S4-b. Triplex 2 Testing with a Dilution Series of Omicron Synthetic RNA (Twist)
###### Chart: Dx Triplex: ORF1ab
| Category | NTC | 50 copies | 50 copies | 500 copies | 500 copies | 5,000 copies | 5,000 copies | 50,000 copies | 50,000 copies |
|---|---|---|---|---|---|---|---|---|---|
###### Chart: Dx Triplex: Spike
| Category | NTC | 50 copies | 50 copies | 500 copies | 500 copies | 5,000 copies | 5,000 copies | 50,000 copies | 50,000 copies |
|---|---|---|---|---|---|---|---|---|---|(Additional results for assays other than S:69/70del are shown here)
62°C:
54°C:
###### Chart: Triplex 2 S:484K
| Category | Well 1 amber | Well 2 amber | Well 3 amber | Well 4 amber | Well 5 amber | Well 6 amber | Well 7 amber | Well 8 amber | Well 9 amber |
|---|---|---|---|---|---|---|---|---|---|
###### Chart: Triplex 2 S:215G
| Category | Well 1 red | Well 2 red | Well 3 red | Well 4 red | Well 5 red | Well 6 red | Well 7 red | Well 8 red | Well 9 red |
|---|---|---|---|---|---|---|---|---|---|
###### Chart: Triplex 2 S:484K
| Category | NTC | 50 copies | 50 copies | 500 copies | 500 copies | 5,000 copies | 5,000 copies | 50,000 copies | 50,000 copies |
|---|---|---|---|---|---|---|---|---|---|
###### Chart: Triplex 2 S:215G
| Category | NTC | 50 copies | 50 copies | 500 copies | 500 copies | 5,000 copies | 5,000 copies | 50,000 copies | 50,000 copies |
|---|---|---|---|---|---|---|---|---|---|

### Slide 6
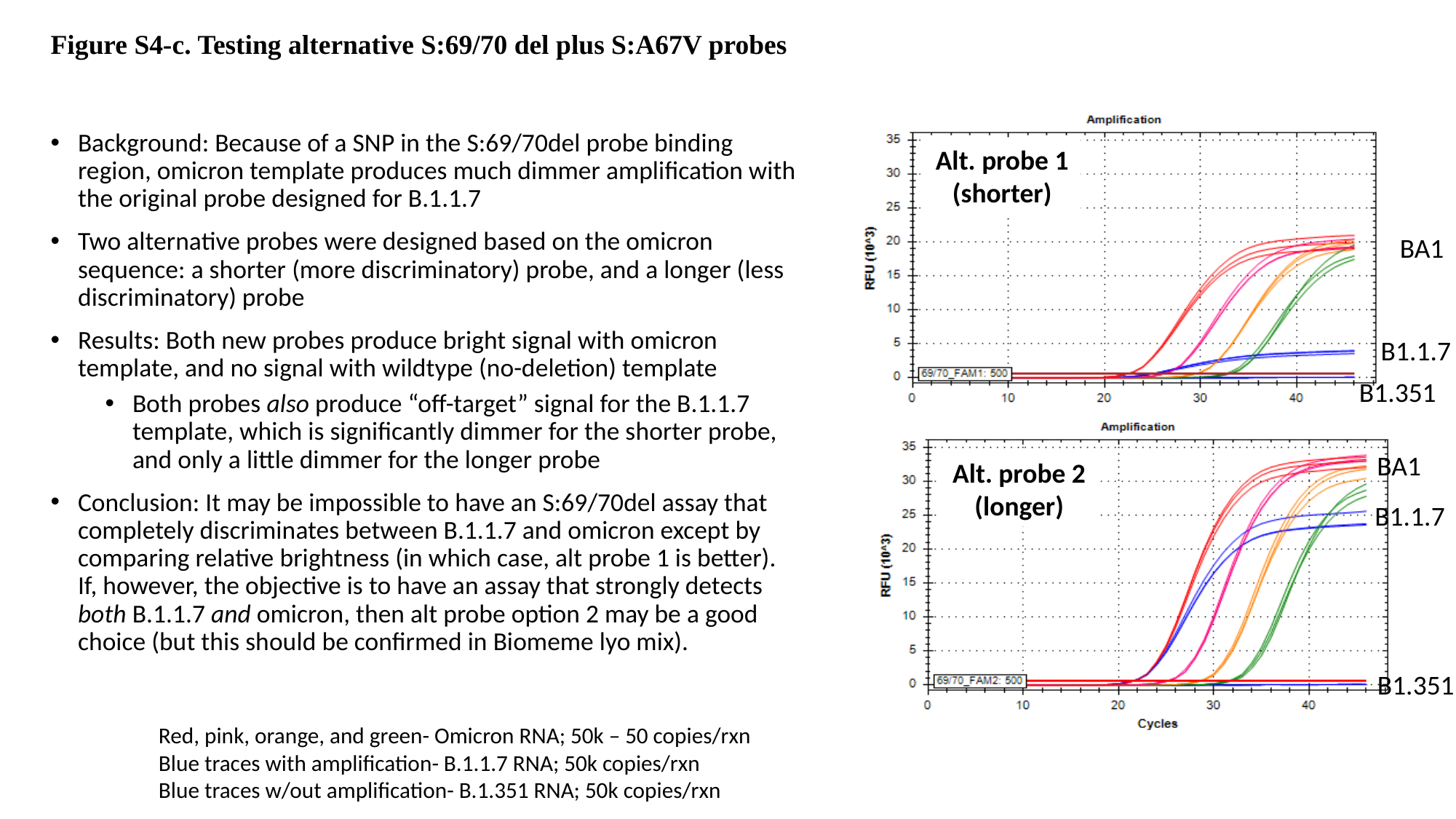

#### Figure S4-c. Testing alternative S:69/70 del plus S:A67V probes
Background: Because of a SNP in the S:69/70del probe binding region, omicron template produces much dimmer amplification with the original probe designed for B.1.1.7
Two alternative probes were designed based on the omicron sequence: a shorter (more discriminatory) probe, and a longer (less discriminatory) probe
Results: Both new probes produce bright signal with omicron template, and no signal with wildtype (no-deletion) template
Both probes also produce “off-target” signal for the B.1.1.7 template, which is significantly dimmer for the shorter probe, and only a little dimmer for the longer probe
Conclusion: It may be impossible to have an S:69/70del assay that completely discriminates between B.1.1.7 and omicron except by comparing relative brightness (in which case, alt probe 1 is better). If, however, the objective is to have an assay that strongly detects both B.1.1.7 and omicron, then alt probe option 2 may be a good choice (but this should be confirmed in Biomeme lyo mix).
Alt. probe 1 (shorter)
BA1
B1.1.7
B1.351
BA1
Alt. probe 2 (longer)
B1.1.7
B1.351
Red, pink, orange, and green- Omicron RNA; 50k – 50 copies/rxn
Blue traces with amplification- B.1.1.7 RNA; 50k copies/rxn
Blue traces w/out amplification- B.1.351 RNA; 50k copies/rxn

### Slide 7
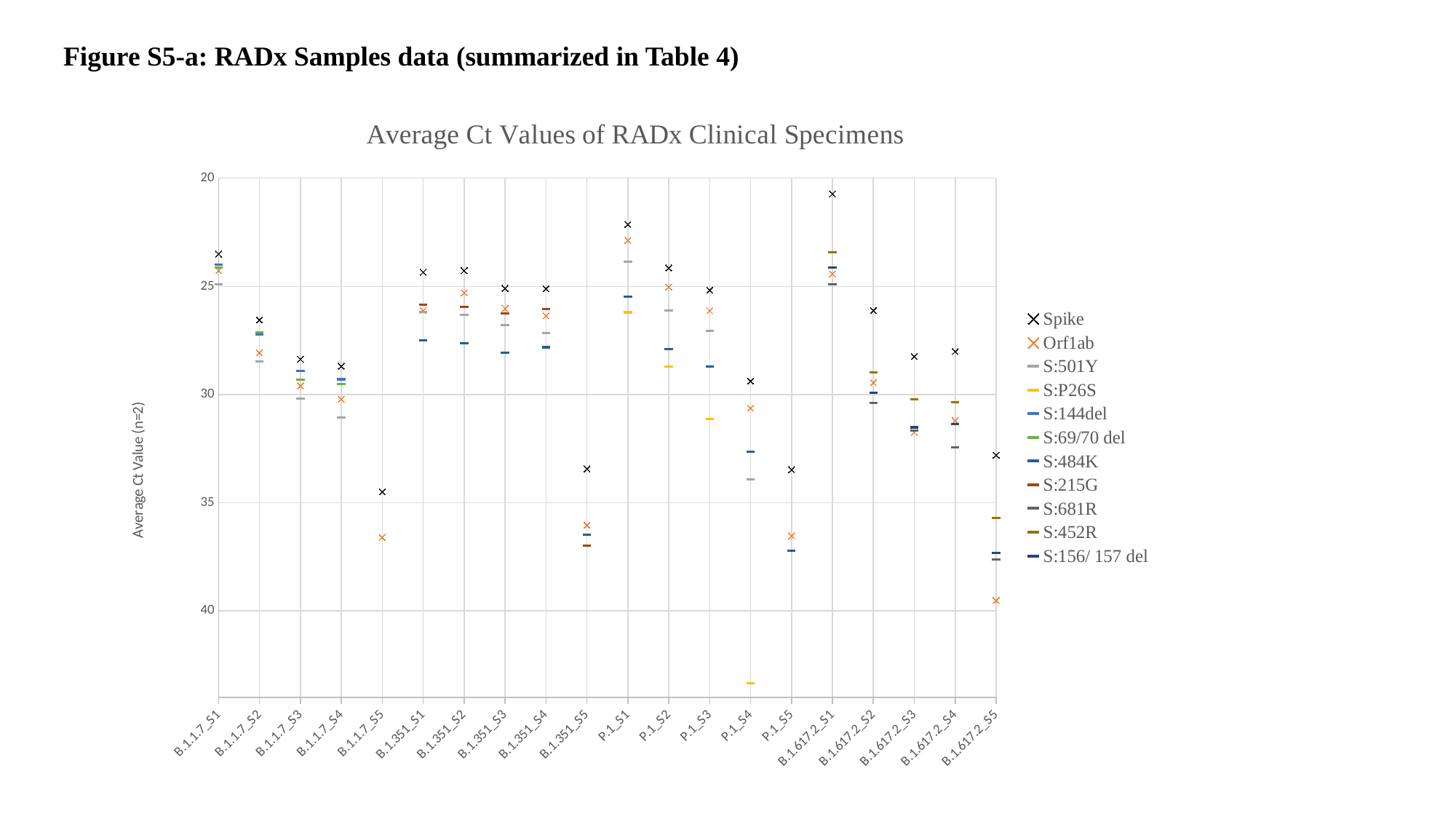

Figure S5-a: RADx Samples data (summarized in Table 4)
###### Chart: Average Ct Values of RADx Clinical Specimens
| Category | Spike | Orf1ab | S:501Y | S:P26S | S:144del | S:69/70 del | S:484K | S:215G | S:681R | S:452R | S:156/ 157 del |
|---|---|---|---|---|---|---|---|---|---|---|---|
| B.1.1.7_S1 | 23.505 | 24.275 | 24.909999999999997 | None | 23.98 | 24.145 | None | None | None | None | None |
| B.1.1.7_S2 | 26.545 | 28.064999999999998 | 28.48 | None | 27.21 | 27.134999999999998 | None | None | None | None | None |
| B.1.1.7_S3 | 28.365000000000002 | 29.59 | 30.195 | None | 28.895 | 29.325 | None | None | None | None | None |
| B.1.1.7_S4 | 28.69 | 30.215000000000003 | 31.055 | None | 29.295 | 29.515 | None | None | None | None | None |
| B.1.1.7_S5 | 34.495000000000005 | 36.6 | None | None | None | None | None | None | None | None | None |
| B.1.351_S1 | 24.345 | 26.115000000000002 | 26.189999999999998 | None | None | None | 27.48 | 25.85 | None | None | None |
| B.1.351_S2 | 24.27 | 25.299999999999997 | 26.305 | None | None | None | 27.630000000000003 | 25.935000000000002 | None | None | None |
| B.1.351_S3 | 25.09 | 26.02 | 26.78 | None | None | None | 28.075000000000003 | 26.255000000000003 | None | None | None |
| B.1.351_S4 | 25.1 | 26.36 | 27.15 | None | None | None | 27.814999999999998 | 26.035 | None | None | None |
| B.1.351_S5 | 33.435 | 36.035 | None | None | None | None | 36.47 | 36.980000000000004 | None | None | None |
| P.1_S1 | 22.134999999999998 | 22.87 | 23.86 | 26.2 | None | None | 25.465 | None | None | None | None |
| P.1_S2 | 24.145000000000003 | 25.03 | 26.1 | 28.715 | None | None | 27.895000000000003 | None | None | None | None |
| P.1_S3 | 25.17 | 26.125 | 27.07 | 31.135 | None | None | 28.715 | None | None | None | None |
| P.1_S4 | 29.369999999999997 | 30.630000000000003 | 33.93 | 43.335 | None | None | 32.65 | None | None | None | None |
| P.1_S5 | 33.475 | 36.53 | None | None | None | None | 37.230000000000004 | None | None | None | None |
| B.1.617.2_S1 | 20.73 | 24.43 | None | None | None | None | None | None | 24.895000000000003 | 23.415 | 24.115000000000002 |
| B.1.617.2_S2 | 26.11 | 29.439999999999998 | None | None | None | None | None | None | 30.395 | 28.965000000000003 | 29.915 |
| B.1.617.2_S3 | 28.235 | 31.75 | None | None | None | None | None | None | 31.66 | 30.235 | 31.515 |
| B.1.617.2_S4 | 28.009999999999998 | 31.195 | None | None | None | None | None | None | 32.445 | 30.345 | 31.35 |
| B.1.617.2_S5 | 32.805 | 39.515 | None | None | None | None | None | None | 37.614999999999995 | 35.715 | 37.325 |

### Slide 8
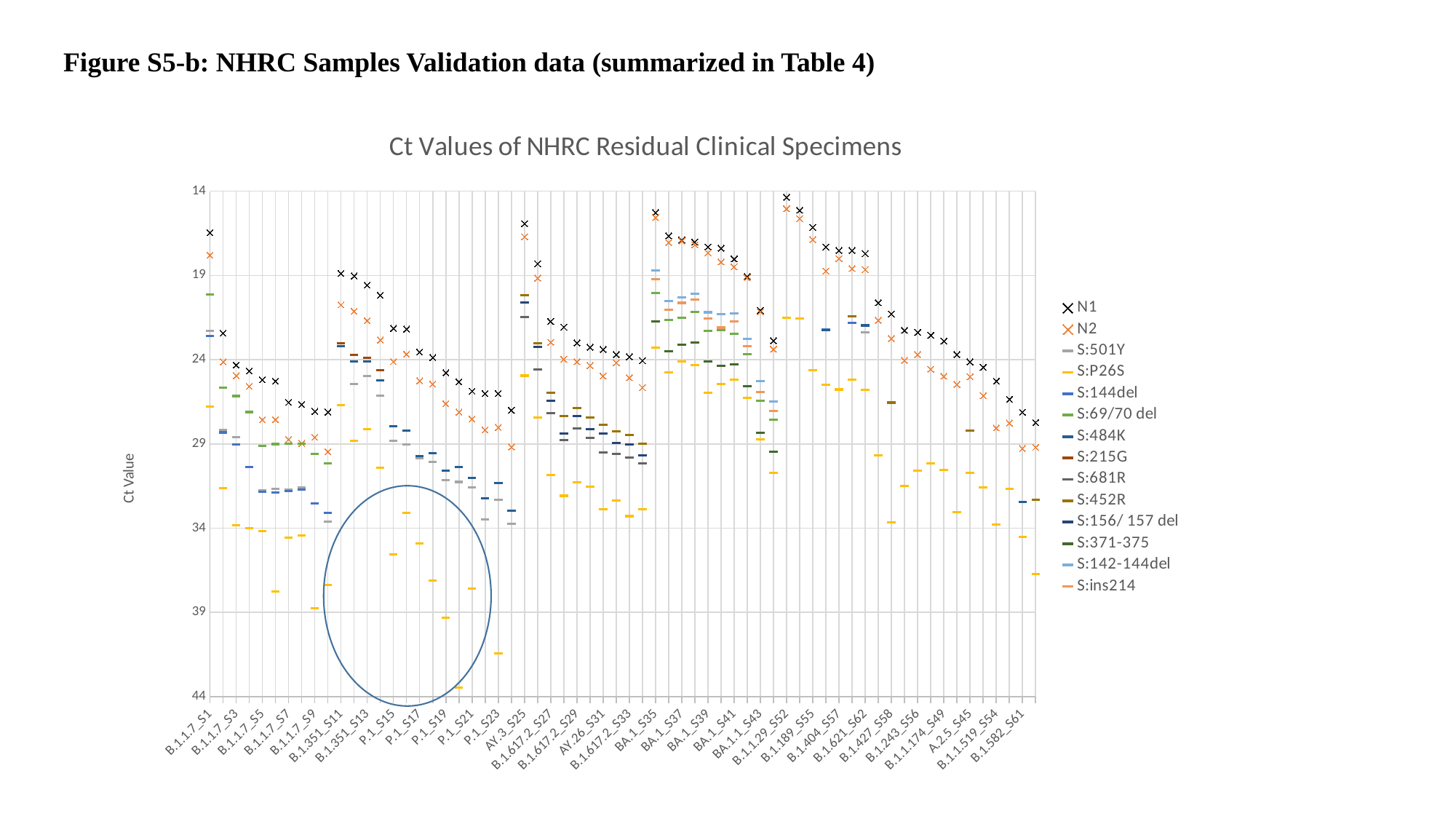

Figure S5-b: NHRC Samples Validation data (summarized in Table 4)
###### Chart: Ct Values of NHRC Residual Clinical Specimens
| Category | N1 | N2 | S:501Y | S:P26S | S:144del | S:69/70 del | S:484K | S:215G | S:681R | S:452R | S:156/ 157 del | S:371-375 | S:142-144del | S:ins214 |
|---|---|---|---|---|---|---|---|---|---|---|---|---|---|---|
| B.1.1.7_S1 | 16.46 | 17.79 | 22.3 | 26.8 | 22.59 | 20.15 | None | None | None | None | None | None | None | None |
| B.1.1.7_S2 | 22.43 | 24.13 | 28.17 | 31.64 | 28.32 | 25.67 | None | None | None | None | None | None | None | None |
| B.1.1.7_S3 | 24.31 | 24.95 | 28.6 | 33.84 | 29.04 | 26.16 | None | None | None | None | None | None | None | None |
| B.1.1.7_S4 | 24.67 | 25.59 | 30.37 | 34.0 | 30.38 | 27.11 | None | None | None | None | None | None | None | None |
| B.1.1.7_S5 | 25.2 | 27.56 | 31.75 | 34.16 | 31.86 | 29.14 | None | None | None | None | None | None | None | None |
| B.1.1.7_S6 | 25.27 | 27.55 | 31.68 | 37.78 | 31.88 | 29.01 | None | None | None | None | None | None | None | None |
| B.1.1.7_S7 | 26.54 | 28.73 | 31.72 | 34.58 | 31.79 | 28.97 | None | None | None | None | None | None | None | None |
| B.1.1.7_S8 | 26.66 | 28.97 | 31.6 | 34.42 | 31.7 | 28.99 | None | None | None | None | None | None | None | None |
| B.1.1.7_S9 | 27.07 | 28.61 | 32.54 | 38.76 | 32.52 | 29.61 | None | None | None | None | None | None | None | None |
| B.1.1.7_S10 | 27.11 | 29.46 | 33.62 | 37.37 | 33.08 | 30.17 | None | None | None | None | None | None | None | None |
| B.1.351_S11 | 18.88 | 20.75 | 23.22 | 26.71 | None | None | 23.18 | 23.04 | None | None | None | None | None | None |
| B.1.351_S12 | 19.03 | 21.12 | 25.43 | 28.81 | None | None | 24.11 | 23.72 | None | None | None | None | None | None |
| B.1.351_S13 | 19.58 | 21.68 | 24.99 | 28.13 | None | None | 24.09 | 23.88 | None | None | None | None | None | None |
| B.1.351_S14 | 20.18 | 22.83 | 26.12 | 30.4 | None | None | 25.25 | 24.63 | None | None | None | None | None | None |
| P.1_S15 | 22.14 | 24.12 | 28.81 | 35.57 | None | None | 27.95 | None | None | None | None | None | None | None |
| P.1_S16 | 22.18 | 23.67 | 29.04 | 33.11 | None | None | 28.21 | None | None | None | None | None | None | None |
| P.1_S17 | 23.55 | 25.25 | 29.84 | 34.92 | None | None | 29.72 | None | None | None | None | None | None | None |
| P.1_S18 | 23.87 | 25.45 | 30.09 | 37.1 | None | None | 29.56 | None | None | None | None | None | None | None |
| P.1_S19 | 24.77 | 26.61 | 31.16 | 39.3 | None | None | 30.58 | None | None | None | None | None | None | None |
| P.1_S20 | 25.31 | 27.11 | 31.26 | 43.46 | None | None | 30.36 | None | None | None | None | None | None | None |
| P.1_S21 | 25.87 | 27.52 | 31.58 | 37.58 | None | None | 31.01 | None | None | None | None | None | None | None |
| P.1_S22 | 26.01 | 28.17 | 33.49 | None | None | None | 32.23 | None | None | None | None | None | None | None |
| P.1_S23 | 26.01 | 28.02 | 32.33 | 41.43 | None | None | 31.31 | None | None | None | None | None | None | None |
| P.1_S24 | 27.0 | 29.18 | 33.73 | None | None | None | 32.95 | None | None | None | None | None | None | None |
| AY.3_S25 | 15.93 | 16.71 | None | 24.95 | None | None | None | None | 21.47 | 20.17 | 20.59 | None | None | None |
| AY.44_S26 | 18.31 | 19.16 | None | 27.45 | None | None | None | None | 24.58 | 23.02 | 23.25 | None | None | None |
| B.1.617.2_S27 | 21.73 | 22.97 | None | 30.85 | None | None | None | None | 27.19 | 25.98 | 26.43 | None | None | None |
| B.1.617.2_S28 | 22.07 | 23.98 | None | 32.08 | None | None | None | None | 28.79 | 27.36 | 28.37 | None | None | None |
| B.1.617.2_S29 | 23.0 | 24.12 | None | 31.3 | None | None | None | None | 28.09 | 26.86 | 27.34 | None | None | None |
| AY.103_S30 | 23.26 | 24.35 | None | 31.56 | None | None | None | None | 28.65 | 27.44 | 28.12 | None | None | None |
| AY.26_S31 | 23.39 | 24.97 | None | 32.9 | None | None | None | None | 29.53 | 27.85 | 28.38 | None | None | None |
| B.1.617.2_S32 | 23.69 | 24.18 | None | 32.35 | None | None | None | None | 29.6 | 28.27 | 28.94 | None | None | None |
| B.1.617.2_S33 | 23.83 | 25.07 | None | 33.29 | None | None | None | None | 29.81 | 28.49 | 29.04 | None | None | None |
| B.1.617.2_S34 | 24.06 | 25.66 | None | 32.87 | None | None | None | None | 30.16 | 28.97 | 29.68 | None | None | None |
| BA.1_S35 | 15.27 | 15.56 | None | 23.29 | None | 20.03 | None | None | None | None | None | 21.73 | 18.7 | 19.21 |
| BA.1_S36 | 16.65 | 17.04 | None | 24.75 | None | 21.65 | None | None | None | None | None | 23.51 | 20.52 | 21.03 |
| BA.1_S37 | 16.9 | 16.95 | None | 24.09 | None | 21.52 | None | None | None | None | None | 23.11 | 20.31 | 20.63 |
| BA.1_S38 | 17.02 | 17.18 | None | 24.33 | None | 21.17 | None | None | None | None | None | 22.97 | 20.1 | 20.45 |
| BA.1_S39 | 17.31 | 17.66 | None | 25.97 | None | 22.28 | None | None | None | None | None | 24.12 | 21.19 | 21.54 |
| BA.1_S40 | 17.38 | 18.2 | None | 25.43 | None | 22.25 | None | None | None | None | None | 24.38 | 21.28 | 22.1 |
| BA.1_S41 | 18.02 | 18.49 | None | 25.2 | None | 22.47 | None | None | None | None | None | 24.26 | 21.24 | 21.72 |
| BA.1_S42 | 19.06 | 19.14 | None | 26.28 | None | 23.68 | None | None | None | None | None | 25.56 | 22.76 | 23.19 |
| BA.1.1_S43 | 21.08 | 21.16 | None | 28.73 | None | 26.43 | None | None | None | None | None | 28.36 | 25.28 | 25.91 |
| BA.1.1_S44 | 22.87 | 23.38 | None | 30.7 | None | 27.57 | None | None | None | None | None | 29.45 | 26.49 | 27.04 |
| B.1.1.29_S52 | 14.36 | 15.04 | None | 21.52 | None | None | None | None | None | None | None | None | None | None |
| B.1.1.139_S47 | 15.13 | 15.63 | None | 21.56 | None | None | None | None | None | None | None | None | None | None |
| B.1.189_S55 | 16.15 | 16.87 | None | 24.61 | None | None | None | None | None | None | None | None | None | None |
| B.1.1.318_S53 | 17.32 | 18.74 | None | 25.48 | 22.22 | None | 22.26 | None | None | None | None | None | None | None |
| B.1.404_S57 | 17.52 | 18.0 | None | 25.77 | None | None | None | None | None | None | None | None | None | None |
| B.1.526.1_S60 | 17.52 | 18.59 | None | 25.18 | 21.83 | None | None | None | None | 21.42 | None | None | None | None |
| B.1.621_S62 | 17.71 | 18.66 | 22.38 | 25.78 | None | None | 21.97 | None | None | None | None | None | None | None |
| B.1.1.162_S48 | 20.62 | 21.66 | None | 29.67 | None | None | None | None | None | None | None | None | None | None |
| B.1.427_S58 | 21.29 | 22.74 | None | 33.65 | None | None | None | None | None | 26.55 | None | None | None | None |
| B.1.1.244_S51 | 22.26 | 24.04 | None | 31.48 | None | None | None | None | None | None | None | None | None | None |
| B.1.243_S56 | 22.38 | 23.69 | None | 30.58 | None | None | None | None | None | None | None | None | None | None |
| B.1_S46 | 22.55 | 24.57 | None | 30.15 | None | None | None | None | None | None | None | None | None | None |
| B.1.1.174_S49 | 22.9 | 24.98 | None | 30.54 | None | None | None | None | None | None | None | None | None | None |
| B.1.628_S63 | 23.69 | 25.47 | None | 33.06 | None | None | None | None | None | None | None | None | None | None |
| A.2.5_S45 | 24.13 | 25.01 | None | 30.72 | None | None | None | None | None | 28.22 | None | None | None | None |
| R.1_S64 | 24.45 | 26.14 | None | 31.59 | None | None | None | None | None | None | None | None | None | None |
| B.1.1.519_S54 | 25.27 | 28.05 | None | 33.79 | None | None | None | None | None | None | None | None | None | None |
| B.1.1.214_S50 | 26.35 | 27.76 | None | 31.67 | None | None | None | None | None | None | None | None | None | None |
| B.1.582_S61 | 27.12 | 29.27 | None | 34.53 | None | None | 32.44 | None | None | None | None | None | None | None |
| B.1.429_S59 | 27.73 | 29.2 | None | 36.71 | None | None | None | None | None | 32.33 | None | None | None | None |
